# Cysteine supplementation reverses immune dysfunction in cancer patients with severe COVID-19

**DOI:** 10.64898/2026.09.21.26363550

**Authors:** Tiffany Merlinsky, Jahan Rahman, William T. Johnson, Lisa McGary, Simon Grassmann, Jennifer Zhang, Abdulraouf Abdulraouf, Hannah L. Kalvin, Katherine Panageas, Jasmine Nicodemus, Elizabeth Cathcart, Ya-Hui Lin, Kinga K. Hosszu, Mirela Berisa, Olga Lyudovyk, Gilles Salles, Jaap J. Boelens, N. Esther Babady, Junyue Cao, Jedd D. Wolchok, Joseph C. Sun, James Heath, Benjamin Greenbaum, Santosha Vardhana

## Abstract

Severe COVID-19 infection in patients with cancer is characterized by a unique pattern of immunologic dysfunction including muted adaptive immune responses. The molecular drivers of immune dysfunction in cancer patients with severe COVID-19 remain unclear, and therapeutic strategies to overcome immune dysfunction in this setting have not been identified.

We performed integrated proteomic and metabolomic profiling of matched cohorts of cancer and non-cancer patients with or without COVID-19 and identified dysfunctional cysteine metabolism as uniquely associated with severe COVID-19 in cancer patients. Treatment of patients with cancer and steroid-refractory COVID-19 with N-acetylcysteine in a prospective clinical trial (NCT04374461) improved clinical outcomes compared with disease severity-matched hospitalized patients during the period immediately preceding clinical trial initiation. N-AC treatment reduced circulating markers of innate inflammation, decreased severe disease-associated MHC-II low monocytes, and increased circulating CD8+ T cell abundance, activation, and effector differentiation. Mechanistically, N-AC reduced prostaglandin E2-driven interactions between suppressive monocytes and T cells, which we confirmed was sufficient to limit T cell expansion and effector differentiation in a dose- and avidity-dependent fashion. Moreover, N-AC reduced the activity of inhibitory, redox sensitive transcription factors such as KLF6, enabling clonal expansion and effector T cell differentiation. We confirmed these observations in two murine models of severe respiratory viral infection, in which N-AC treatment significantly enhanced lung-infiltrating CD8+ T cell abundance. These findings establish cysteine supplementation as a viable therapeutic strategy to reverse redox-driven immune dysregulation in severe respiratory viral infection, particularly in the high-risk cancer population.

**One Sentence Summary:** Merlinsky et al show that cysteine supplementation can therapeutically reverse immune dysregulation in cancer patients with severe COVID-19 infection.

## INTRODUCTION

Respiratory infections are a significant cause of morbidity in patients with cancer, accounting for upwards of 10% of hospitalizations in cancer patients and nearly a third of hospitalizations in patients with hematologic malignancies(*1*). Patients with cancer admitted to the hospital for pneumonia have a substantially increased risk of mortality and this increased risk persists even when patients are not on active treatment(*2, 3*). This increased risk is exacerbated during severe pandemic infections, most recently during the COVID-19 pandemic mortality rates in hospitalized patients with cancer reached as high as 50%(*4, 5*).

Severe COVID-19 is increasingly appreciated as a disease of immune dysregulation, with excessive innate immune activation as a key predictor of tissue damage and mortality(*6*). As such, the mainstay of therapy for immunocompetent patients with severe viral infections, including COVID-19 is immunosuppression(*7*). Limited available data in cancer patients, however, indicate that immunosuppression is not beneficial and possibly even harmful(*8, 9*). We and others previously reported that defects in adaptive immunity underly both short and long-term morbidity and mortality in COVID-19 infected patients with cancer, suggesting that an adequate adaptive immune response is required for clearance in these high-risk individuals(*10–13*). Consistent with this hypothesis, patients with persistent COVID-19 despite corticosteroid therapy tend to have persistent innate inflammation but often blunted or even muted CD8+ T cell responses(*14, 15*). Neither drivers of immune dysregulation nor validated therapeutic strategies to either reverse pathologic myeloid dysfunction or restore CD8+ T cell immunity in patients with severe respiratory viral infections have been identified.

We proposed that metabolic dysregulation might contribute to immune dysfunction in severe COVID-19, given that severe disease-associated systemic hypoxemia can have profound metabolic consequences that impact the phenotype and function of both innate and adaptive immune cells(*6, 16*). To test this hypothesis, we performed a multi-omic analysis of cancer patients with acute COVID-19 infection and compared them to COVID-19 patients from the well-annotated INCOV dataset(*17, 18*) as well as age-matched control patients without cancer or COVID-19, identifying blunted T cell immunity and impaired cysteine recycling as hallmarks of patients with severe disease. Restoring cysteine availability with N-acetylcysteine (N-AC) showed encouraging clinical outcomes in a single-institution phase II study (NCT04374461). Analysis of both samples from patients obtained during study and mouse models of severe respiratory viral infections indicated that N-AC reversed pathologic accumulation of immunosuppressive monocytes and enabled quantifiable effector CD8+ T cell expansion. We identified redox-sensitive transcription factors associated with cell stress, immunosuppression, and activation underlying the observed changes in myeloid and CD8+ T cell phenotypes. These findings establish redox dysfunction as a key contributor to COVID-19 driven immune dysfunction and nominate cysteine supplementation as a rational strategy to reverse immune dysregulation in patients with severe viral infections.

## RESULTS

### Immune and metabolic dysfunction distinguishes cancer patients with severe COVID-19

To identify systemic immune and metabolic features contributing to severe COVID-19 in cancer patients, we analyzed plasma samples from patients at Memorial Sloan Kettering Cancer Center (MSKCC), including hospitalized patients with acute COVID-19, patients who had recovered from COVID-19 (‘convalescent’), and hospitalized patients who had not tested positive for COVID-19 (‘no COVID’) (**Fig. 1A**). These samples were compared with plasma samples from the INCOV cohort spanning a range of COVID-19 severities, recovered patients, and age and comorbidity-matched controls from unexposed healthy patients (see **Table S1** and Methods). 68% of MSKCC patients had hematologic malignancies, which is consistent with patients with hematologic malignancies having an odds ratio of 2.5 for hospitalization in work previously published from MSKCC(*5*). Moreover, MSKCC and INCOV patients did not differ substantially with respect to age, gender, ethnicity, race, or median days between COVID diagnosis and sample acquisition (**Table S1**).

**Figure 1.**
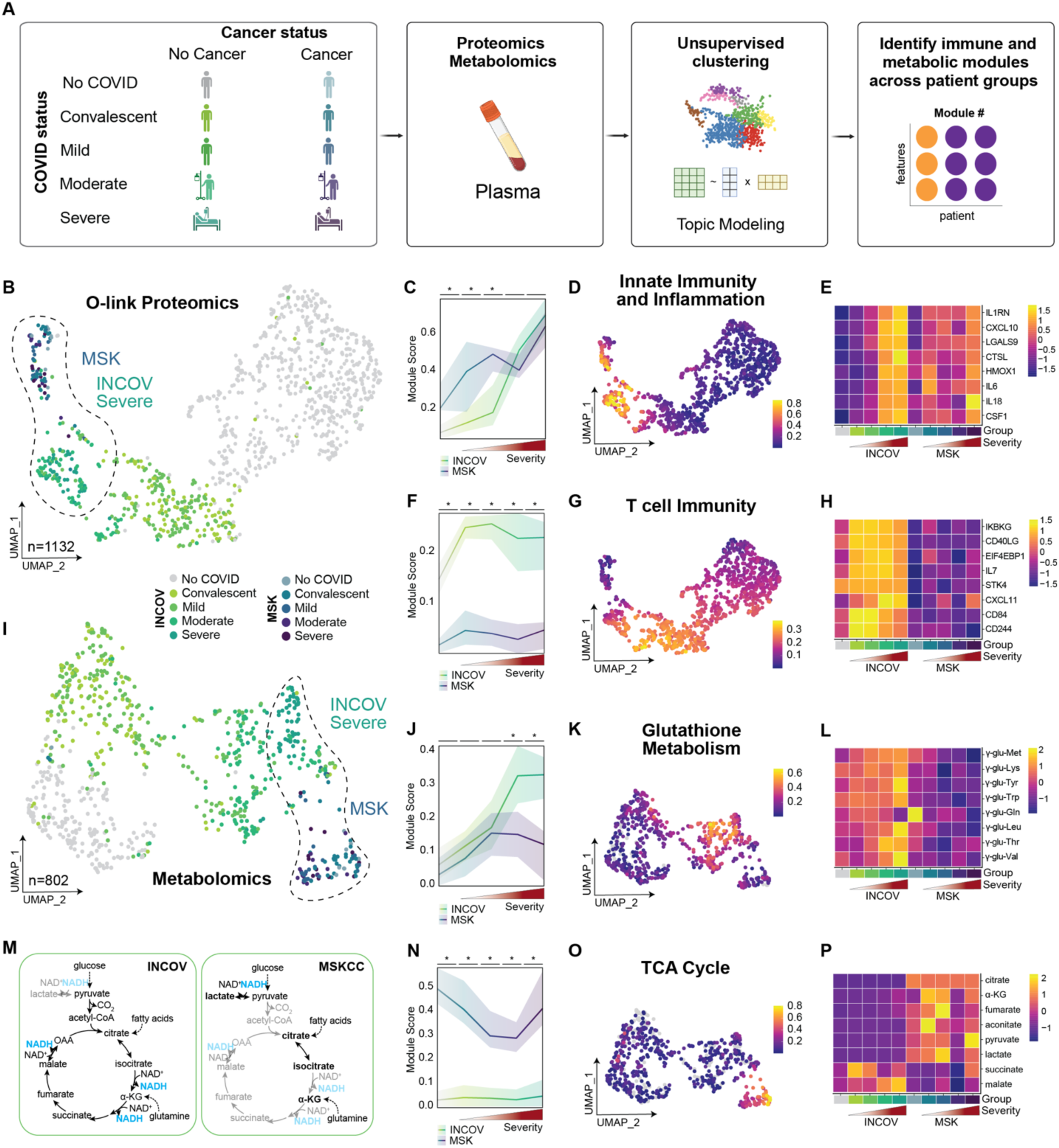
Immune and metabolic dysregulation characterizes cancer patients with severe COVID-19. **(A)** Schematic of sample collection, processing, and multi-omic analysis of plasma samples from patients from cancer (MSK) and cancer-free (INCOV) datasets. **(B)** UMAP of O-link proteomics data. Each sample is represented by a point (n= 1132). **(C,D)** Line graph (C) and UMAP projection (D) of Proteomics module 2 (Innate Immunity and Inflammation) scores calculated by consensus NMF. Average is bolded, and 25^th^ to 75^th^ percentile highlighted. **(E)** Average scaled expression of individual representative proteins from module 2. **(F,G)** Line graph (F) and UMAP projection (G) of Proteomics module 3 (T cell Immunity) scores calculated by consensus NMF. **(H)** Average scaled expression of individual representative proteins from module 3. **(I)** UMAP of Metabolomics data. Each sample is represented by a point (n= 802). **(J,K)** Line graph (J) and UMAP projection (K) of Metabolomics module 2 (Glutathione Metabolism) scores calculated by consensus NMF. **(L)** Average scaled expression of individual representative metabolites from module 2. **(M)** Schematic of TCA cycle dynamics in patients with severe COVID-19 in the INCOV and MSKCC cohorts based on interpretation of data represented in O-Q. **(N,O)** Line graph (N) and UMAP projection (O) of Metabolomics module 4 (TCA Cycle) scores calculated by consensus NMF. **(P)** Average scaled expression of individual representative metabolites from module 4. *p<0.001 by pairwise Mann-Whitney U after adjustment for multiple comparisons by Bonferroni correction (C,F,J,N).

We employed non-negative matrix factorization (NMF) to uncover latent modules in our plasma protein and metabolic datasets(*19*). Unlike conventional pathway enrichment analyses, which rely on predefined reference sets and lack directionality, NMF is an unsupervised machine learning approach that captures coordinated patterns among proteins and metabolites, including those without standard database annotations (e.g. KEGG, HMDB). This enabled us to detect functionally coherent modules that would be overlooked by traditional methods due to missing identifiers or incomplete pathway mappings. Moreover, the modules exposed interpretable biological bottlenecks and helped us distinguish between adaptive versus maladaptive processes.

NMF of plasma proteomic data generated four distinct modules (**Fig. 1B, S1A and Table S2**). Modules 2 and 3 revealed striking differences between patients with and without cancer (**Table S3**). Module 2, enriched for proteins involved in inflammatory signaling and monocyte activation, increased with COVID severity in both cohorts, but was elevated in MSKCC patients at baseline and with mild or convalescent disease (**Fig. 1C-E**). This module is consistent with published data showing an association of innate inflammation with COVID-19 disease severity (*20*) and suggests that this association is intact in the cancer population. In contrast, Module 3, comprising proteins involved in T cell recruitment, activation, and proliferation, was robustly induced in patients without cancer, but was blunted or even absent in patients with cancer, suggesting that impaired T cell immunity may contribute to disease severity specifically in the cancer population, as previously shown in patients with cancer(*10*) (**Fig. 1F-H**). The remaining modules highlighted programs with shared kinetics between MSKCC and INCOV patients, including a COVID-19 severity module associated with tissue remodeling (SPON2, FIGF, TEK, THBS2) and protection against oxidative stress (SOD2, HSPB1) (**Fig. S1A**).

Next, we applied NMF to plasma metabolite data and identified seven distinct modules (**Fig. 1I, S1B and Table S4**). Module 2 was primarily defined by g-glutamylated amino acids (**Fig. 1J-L**), which are extracellular products of the glutathione cycle. As most cells are unable to import intact glutathione (L-γ-glutamyl-L-cysteinyl-glycine), extracellular glutathione it is broken down into a g-glutamyl moiety and cysteinyl-glycine by γ-glutamyl transferase (*GGT*); the cysteinyl-glycine provides rate-limiting amino acid cysteine for intracellular glutathione synthesis while the g-glutamyl group combines with acceptor amino acids via GGT generating g-glutamyl amino acids(*21*). Systemic accumulation of g-glutamyl amino acids thus reflects increased demand for glutathione; consistent with this, *GGT* is a well-established prognostic marker in conditions driven by oxidative stress, such as acute myocardial infarction or pulmonary embolism(*22, 23*). Expression of Module 2 increased with COVID-severity in INCOV (**Fig. 1J-L and Table S5**), consistent with published data showing increased expression of genes related to glutathione metabolism in immune cells from patients with severe COVID-19(*24*). Interestingly, Module 2 expression was blunted with COVID-severity in MSKCC, suggesting an inability to activate *GGT*-dependent cysteine regeneration in these patients.

Cysteine is a critical amino acid for the biosynthesis of redox-sensitive molecules, including iron-sulfur cluster proteins required for mitochondrial electron transport. Accordingly, cysteine limitation has been associated with impaired mitochondrial tricarboxylic acid (TCA) cycle activity(*25*). We observed an enrichment of Module 4, containing several central carbon metabolites, within the plasma of MSKCC patients (**Table S5**), including increased accumulation of pyruvate, lactate and upstream TCA cycle metabolites citrate, aconitate, alpha ketoglutarate, and fumarate and decreased accumulation of downstream metabolites succinate and malate. Pyruvate, citrate, and alpha ketoglutarate are all metabolized by NAD+ dependent dehydrogenases; the extracellular accumulation of these central carbon intermediates is consistent with impaired TCA cycle oxidation of either glucose, fatty acid, or glutamine-derived carbons (**Fig. 1M-P**) and was recently observed in mice lacking endogenous cysteine synthesis who were fed a cysteine-deficient diet(*26*). The remaining modules captured metabolic programs associated with lipid, nucleotide and amino acid metabolism and did not substantially differ between the INCOV and MSKCC cohorts, although some correlated with disease severity (**Fig. S1B** and **Table S5**).

### N-AC improves clinical outcomes in cancer patients with severe COVID-19

Our laboratory previously showed that cysteine supplementation could reverse mitochondrial dysfunction in exhausted CD8+ T cells and restore their anti-tumor efficacy (*27*). To test whether replenishing cysteine could improve outcomes in cancer patients with severe COVID-19, we rapidly initiated an investigator-initiated, two-arm, phase II study of N-acetylcysteine (N-AC) in patients with severe COVID-19 who were hospitalized at MSKCC (NCT04374461) (**Fig. 2A**).

**Figure 2.**
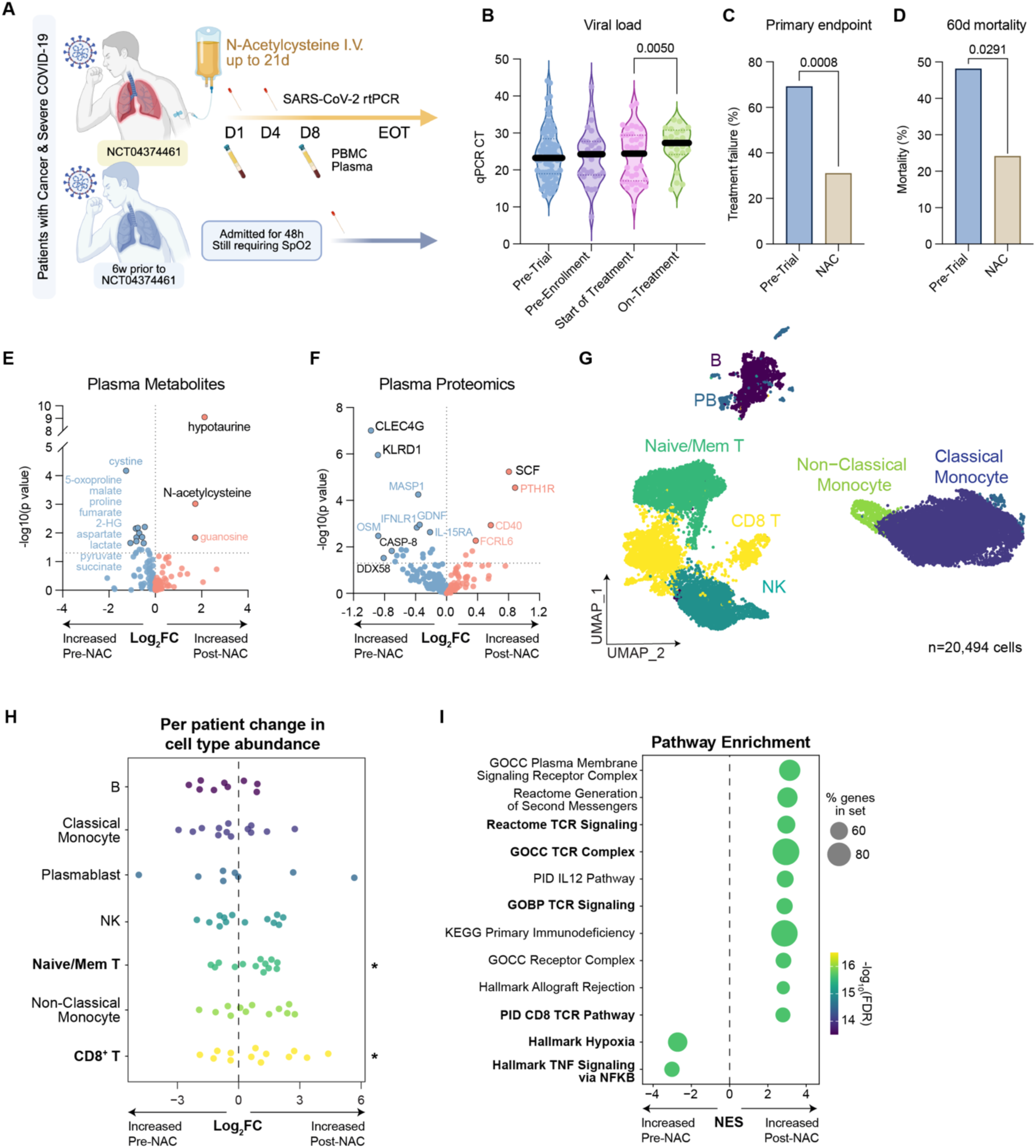
Systemic N-Acetylcysteine administration improves clinical outcomes and reverses hallmarks of immune and metabolic dysfunction in cancer patients with severe COVID-19. **(A)** Schematic of sample collection from hospitalized patients enrolled in NCT04374461 and hospitalized patients who would have met criteria for study enrollment in the 6 weeks prior to study opening (10 March 2020 to 7 April 2020). **(B)** Median cycles required for viral detection from nasopharyngeal swabs obtained from patients at indicated timepoints on study, compared with patients from the pre-trial period. (C,D) Frequency of failure to achieve study endpoint (C) and mortality (D) in patients on NCT04374461 compared with patients from the pre-trial period who would have met criteria for study enrollment. (E,F) Change in abundance of plasma metabolite (E) and plasma proteins from O-link Inflammation and Inflammatory Response panels (184 total proteins) (F) following treatment initiation in 23 patients with paired pre-and on-treatment plasma samples. **(G)** UMAP projection of peripheral blood immune cells (n= 20,494 cells) isolated from 13 paired pre- and on-treatment samples from patients on NCT04374461, based on weighted transcriptomic and cell surface protein profiles. **(H,I)** Relative change in cell type abundance per patient (H) and enrichment of preranked genesets (I) following initiation of N-AC treatment. P values determined by unpaired t-test (pre-trial vs. pre-enrollment or start of treatment) or paired t-test (start of treatment vs. on-treatment) (B) and Fisher’s Exact Test (C,D). Significant results from scCODA analysis are shown in bold with an asterisk (H).

Full details of the protocol can be found in the **Supplementary Methods.** Briefly, patients were eligible if they were hospitalized at MSKCC for COVID-19 and had severe disease (defined as requiring Supplementary oxygen of 2L or more by nasal cannula to maintain an SpO2 of 95%). Patients were assigned to different arms based on whether they required ICU-level care or mechanical ventilation (Arm A) or not (Arm B). Patient characteristics across both Arms are in **Table S6**. A total of 42 patients were enrolled and included in the final efficacy analysis, including 13 (31%) patients on arm A and 29 (69%) patients on arm B. The patient population was heavily pre-treated; the median time from hospitalization to treatment initiation was 7.5 days (range, 2-47 days). All but one of the 42 patients had received some form of anti-COVID supportive care therapy at the time of enrollment, including 39 (93%) patients treated with steroids, 33 (79%) patients treated with remdesivir, 29 (69%) patients treated with monoclonal antibodies or convalescent plasma, and 23 (55%) who received all three modalities. All patients required at least one positive SARS-CoV-2 RNA sample prior to enrollment, of whom forty (95%) had quantitative PCR performed for estimation of viral load (**Fig. 2B**). Twenty-two patients (52%) had at least one additional SARS-CoV-2 RNA sample either while on protocol or after coming off protocol. In patients who had samples obtained both prior to and following N-AC treatment, the number of cycles required to detect SARS-CoV-2 RNA significantly increased with treatment. In contrast, the number of cycles required for detection did not differ between hospital admission and study enrollment, suggesting that N-AC treatment was associated with an improvement in SARS-CoV-2 viral load (**Fig. 2B**).

The primary endpoint of this study was clinical improvement, defined as absence of treatment-related toxicity or death (Arm A) or absence of ICU transfer, intubation, treatment toxicity, or death (Arm B). In total, 23 (55%) patients met the predetermined clinical definition of treatment success, including 20 of 29 patients on Arm B (69%) (**Fig. 2C-D**). As this was a non-randomized study in which all patients received treatment, we compared our study population to published outcomes of hospitalized patients at MSKCC from the six weeks immediately preceding study initiation who would have met criteria for study inclusion on Arm B (**Fig. 2A, S2A, and Table S6**) (*5*). Patients from the pre-trial period who met criteria for study inclusion did not differ from enrolled patients with respect to age, gender, race, malignancy subtype, frequency of active treatment, frequency of B cell depleting therapy, or standard treatment for COVID-19, including steroids, remdesivir, or monoclonal antibodies (**Table S6**). Moreover, patients from the pre-trial period who met criteria for study inclusion had a similar estimated viral load to enrolled patients prior to treatment initiation (**Fig. 2B**). Despite this, patients who received treatment on study had lower rates of both the primary endpoint (**Fig. 2C**) and all-cause mortality (**Fig. 2D**) compared with patients from the pre-trial period. Of note, the treatment success of conventional therapies in the pre-trial period was substantially worse (30%) than estimated at time of study design (60%); as such these results are consistent with N-AC as a promising therapeutic strategy for steroid-refractory COVID-19 despite not meeting the pre-specified threshold of success (80%).

### N-AC reverses COVID-associated metabolic dysfunction and remodels systemic immunity

To understand the mechanism by which N-AC improved clinical outcomes in patients with severe COVID-19, we first assessed the impact of N-AC on systemic metabolism. We performed an unbiased metabolomic analysis of plasma samples from 23 patients prior to and following initiation of treatment (n=59). Unbiased analysis of 154 metabolites revealed increased levels of N-AC and hypotaurine with a reciprocal decrease in 5-oxoproline, indicating enhanced cysteine-dependent biosynthesis and reduced dependence on cysteine recycling (**Fig. 2E** and **Table S7**). In addition, N-AC treatment decreased accumulation of metabolites associated with mitochondrial ETC dysfunction, including lactate, 2-hydroxyglutarate, and TCA cycle metabolites (**Fig. 2E** and **Table S7**), suggesting that cysteine supplementation with N-AC may support mitochondrial function in patients with severe COVID-19.

Next, we determined the impact of N-AC treatment on inflammation and immunity. Using the O-link proteomic platform, we analyzed plasma abundances of 184 proteins from 23 paired pre-and on-treatment plasma samples (n=46) (**Fig. 2F** and **Table S8**). N-AC decreased levels of proteins associated with chronic innate inflammation and adaptive immune suppression, including CLEC4G, which can induce myeloid hyperactivation by binding SARS-CoV-2 spike protein or directly suppress CD8+ T cell activation (*28, 29*). To further our understanding of how N-AC treatment shapes the immune landscape, we performed single cell multi-omic profiling of peripheral blood mononuclear cells (PBMC) from 13 paired pre- and on-treatment samples (n=26) (**Fig. 2G and S2B-F**). We identified 7 broad immune lineages with expression of cell-type specific markers: B cells, plasmablasts, naïve/memory T cells, CD8+ T cells, NK cells, non-classical monocytes, and classical monocytes.

We next asked whether N-AC treatment was associated with changes in the abundance in any of these cell types. To do so, we employed scCODA(*30*), which uses a Bayesian model to address data sparsity and cell number heterogeneity in single cell datasets and visualized the change in cell type abundance using a generalized linear model (GLM) with a quasibinomial family, which models the log-odds of cell type abundance while accounting for variability in total cell numbers per sample. This approach was specifically developed for use when per-sample cell counts vary, and when some clusters may have low representation across samples to protect from artificial inflation and has been used in recently established methods such as Milo(*31*) (**Fig. S2G,H**). N-AC treatment led to a marked expansion of both naïve/memory T cells and CD8+ T cells by scCODA (**Fig. 2H** and **Table S9**). These changes were also present in the raw per-patient data; for example, treatment was associated with a 35% increase in CD8+ T cells (14.4% to 19.5%) and a 28% increase in naïve/memory T cells (14.7% to 18.8%) (**Fig. S2I** and **Table S9**). Further, gene set enrichment analysis highlighted increased expression of genes associated with TCR signaling (**Fig. 2I**), which were primarily expressed by T cells (**Fig. S2J,K**) as well as decreased expression of genes associated with Hypoxia and TNF signaling (**Fig. 2I**), which were enriched primarily within monocytes (**Fig. S2J,L**). Importantly, these changes were seen in both patients who did and did not meet the primary endpoint, suggesting that the immunological changes seen following N-AC therapy did not merely reflect clinical improvement (**Fig. S2M,N**). Together, these data indicate that systemic administration of N-AC restores mitochondrial function and remodels systemic immune responses in patients with severe COVID-19.

### N-AC reprograms suppressive severity-associated monocytes

Dysfunctional (CD11b+MHC-II^low^) monocytes are the single largest immunological predictor of COVID-19 disease severity as well as the most robust predictor of dexamethasone treatment failure(*32*). To explore N-AC driven alterations in monocyte phenotypes, we subclustered peripheral blood monocytes and identified 8 subpopulations (**Fig. 3A and S3A**). N-AC treatment was associated with a decrease in MHC-II^low^ classical monocytes expressing *HIF1A* and *AREG*, the latter of which has been associated with pulmonary fibrosis and pathologic lung remodeling in COVID-19 (**Fig. 3B and Table S10**)(*33*). These changes were also present in the raw per-patient data; for example, treatment was associated with statistically significant decreases in classical HIF1A+AREG+ monocytes (23.8% to 9.1%) and granulocytic HIF1A+AREG+ monocytes (16.3% to 10.5%) as well as a significant increase in non-classical monocytes (5.7% to 9.3%) (**Fig. S3B**). DEG analysis showed treatment-dependent decreased expression of oxidative stress response genes, including *HIF1A*, metallothioneins (e.g. *MT2A, MT1E*), *AREG*, *TNXIP*, and increased expression of genes encoding MHC-II (e.g. *HLA-DRB1, HLA-DRA, HLA-DPB1*) (**Fig. 3C** and **Table S11**). Accordingly, gene set enrichment analysis revealed significant upregulation of antigen processing and presentation pathways and increased translational activity by way of decreased expression of stress-associated ribosomal pathways (**Fig. 3D-E and S3C**).

**Figure 3.**
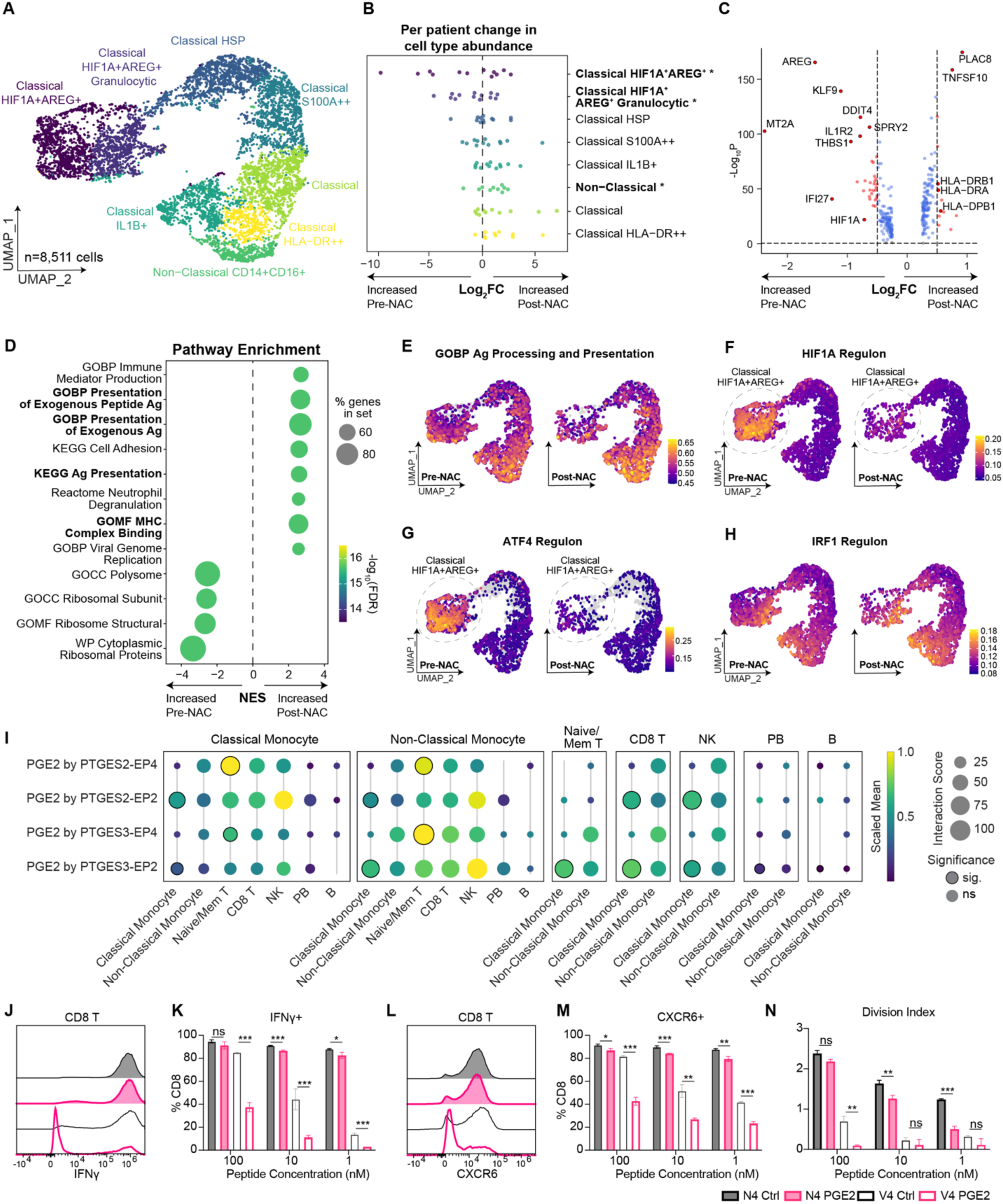
N-Acetylcysteine reprograms MHC-II^low^ monocytes. **(A)** UMAP of monocytes (n= 8,511 cells), based on weighted transcriptomic and gene regulatory network profiles inferred by SCENIC. **(B)** Relative change in cell type abundances per patient in pre- versus on-treatment samples. **(C,D)** Volcano plot of differentially expressed genes (C) and change in preranked geneset enrichment (D) in monocytes identified by pseudobulk analysis of pre-versus on-treatment samples. **(E)** Feature plot depicting expression of GOBP Processing and Presentation of Exogenous Peptide Antigen pathway normalized enrichment scores (Enrichr) in pre- and on-treatment monocytes. **(F-H)** Feature plots depicting expression of HIF1A (F), ATF4 (G), and IRF1 (H) regulon activity (AUCell) in monocytes from pre- and on-treatment samples as determined by SCENIC. **(I)** Dot plot depicting scaled mean expression of Prostaglandin E2 interacting partners per cell-type pair in pre-treatment PBMCs using CellPhoneDBv5. **(J-N)** Immunophenotyping of OT-I CD8+ T cells 4 days after co-culture with antigen-pulsed dendritic cells in the presence or absence of exogenous Prostaglandin E2. **(J)** Representative histograms of IFNg expression in T cells co-cultured with 100 nM peptide pulsed DCs. **(K)** Quantification of IFNg positivity in stimulated CD8+ T cells. **(L)** Representative histograms of CXCR6 expression in T cells co-cultured with 100 nM peptide pulsed DCs. **(M)** Quantification of CXCR6 positivity in stimulated CD8+ T cells. **(N)** Division index of CD8+ T cells determined by Cell Trace Violet labeling. *p<.05 **p<.01 ***p<.001. P values determined by unpaired t-test (K,M,N). Significant results from scCODA analysis are shown in bold with an asterisk (B).

We next sought to identify transcription factors driving NAC-dependent monocyte cell state changes. We used SCENIC to create a transcription factor activity matrix, based on transcription factor target gene co-expression networks and regulatory motif analysis from single cell transcriptomics data whose activity is quantified based on gene expression rankings on a per cell basis (AUCell) (*34, 35*). Using this readout, the activity of transcription factors ATF4 and HIF1A emerged as highly sensitive to NAC-treatment and were most notably decreased in the HIF1A+AREG+ monocyte clusters (**Fig. 3F-G and Fig. S3D,E**). Additionally, increased activity of IRF1, a master regulator of MHC-II genes, was observed across multiple monocyte clusters following treatment, suggesting coordinated expansion of existing MHC-II^hi^ populations and reprogramming of suppressive MHC-II^low^ populations to enhance antigen presentation capacity (**Fig. 3H and Fig. S3F**).

### N-AC reverses suppressive monocyte-T cell interactions and promotes effector CD8+ T cell expansion

Next, given our observations of increased antigen presentation in monocytes with N-AC by gene, pathway, and transcription factor analysis, we asked whether N-AC alters monocyte interactions with other cell types. To test this, we utilized CellPhoneDBv5 to identify and score ligand-receptor interactions from single-cell transcriptomics data(*36*). As expected, the dominant interactions on treatment were activating and associated with productive T cell priming (ICAM2, ICAM3, SEMA4D_CD45, CD47_SIRPG, SELPLG_SELL) (**Fig. S3G and Table S12**). In contrast, the dominant interactions pre-treatment were inhibitory, including multiple interacting pairs involving myeloid cell-derived prostaglandin E2 (PGE2) and PGE2 receptors from naïve and memory T cells (**Fig. 3I and Table S13**). Elevated PGE2 has been reported in COVID-19 with serum concentrations of 10-30 ng/mL in severe disease(*37*). Furthermore, PGE2 has been shown to inhibit CD8+ T cell expansion and effector function by compromising mitochondrial function(*38, 39*), but the impact of PGE2 on T cell priming remains unclear, and whether the effect of PGE2 is uniform across antigen density and affinity has not been well established. To determine how myeloid cell-derived PGE2 might alter T cell priming in severe COVID-19, we co-cultured naïve CellTrace Violet-labeled OT-I transgenic T cells and dendritic cells pulsed with peptides of either a high (SIINFEKL, N4) or low (SIIVFEKL, V4) affinity for the OT-I TCR (**Fig. S3H,I**)(*40*). We found that T cell priming was highly sensitive to PGE2 at doses as low as 1 ng/mL when stimulated by antigens with low avidity or at low abundance, and escalating PGE2 doses amplified these effects, indicating that elevated PGE2 within a range seen in COVID-19 patients can impact T cell priming (**Fig. 3J-N and S3J**). Interestingly, while the effect of PGE2 on high avidity T cells was seen primarily at the level of proliferative capacity (as reflected by the decrease in division index), PGE2 markedly suppressed not only the proliferative capacity but also expression of both activation markers (CD44+, CD25+CD69+) and markers of cytotoxic function (CXCR6, IFN-g) in OT-I T cells responding to a low avidity peptide (**Fig. 3J-N and S3K**). While SPR-based monomeric affinity of SIINFEKL for OT-I TCR (Kd 2-5 uM) is at the high end of established immunodominant COVID-19-derived peptides on HLA-A2*02:01 (2-50 uM), the functional avidity of SIINFEKL for OT-I (EC50 of 10 pM) is multiple orders of magnitude higher than the functional avidity of most viral or microbial antigens, including established immunodominant COVID-19-derived peptides on HLA-A2*02:01 (EC50 of 0.1-3 uM)(*40–42*), making the effect of PGE2 on T cells stimulated by ‘low avidity’ peptides such as SIIVFEKL, which has an EC50 of 10 nM, a more accurate reflection of the impact of PGE2 might have on true COVID-19 peptide-dependent T cell immunity.

These results are suggestive of redox-dependent suppression of T cell priming by dysfunctional monocytes in COVID-19. Decreased circulating T cells and TCR signaling are associated with increased COVID-19 severity, including in patients with cancer(*10, 43*). Given that cancer patients displayed a muted adaptive immune response (**Fig. 1G-I**) and our observed improved antigen presentation capacity of myeloid cells following N-AC treatment (**Fig. 3C,D**), we next investigated the impact of N-AC on T cell immunity. Clustering of CD8+ T cells revealed 9 subpopulations (**Fig. 4A and S4A-C**), including a novel suppressed naïve T cell subset (Naïve PELI1+) marked by expression of negative regulators of T cell activation (*PELI1, TOB1, JUND, FOXP1*)(*44–46*), in addition to canonical naive T cell markers (*IL7R, LTB, TCF7, LEF1*, CD45RA+, CD45RO-) (**Fig. 4B and S4A**). This subset decreased significantly in abundance following N-AC treatment (33.9% to 20.9%), suggesting that the Naïve PELI1+ state is redox sensitive (**Fig. 4C, S4D** and **Table S14**). Moreover, expression of *PELI1*, which encodes for a ubiquitin ligase that negatively regulates T cell activation (*44*), decreased both overall and specifically within this cluster following N-AC treatment (**Fig. 4D, Fig. S4E, and Table S15**). We confirmed that activation of primary human T cells under hypoxia was sufficient to induce PELI1 expression in a manner that could be reversed by N-AC (**Fig. S4F**).

**Figure 4.**
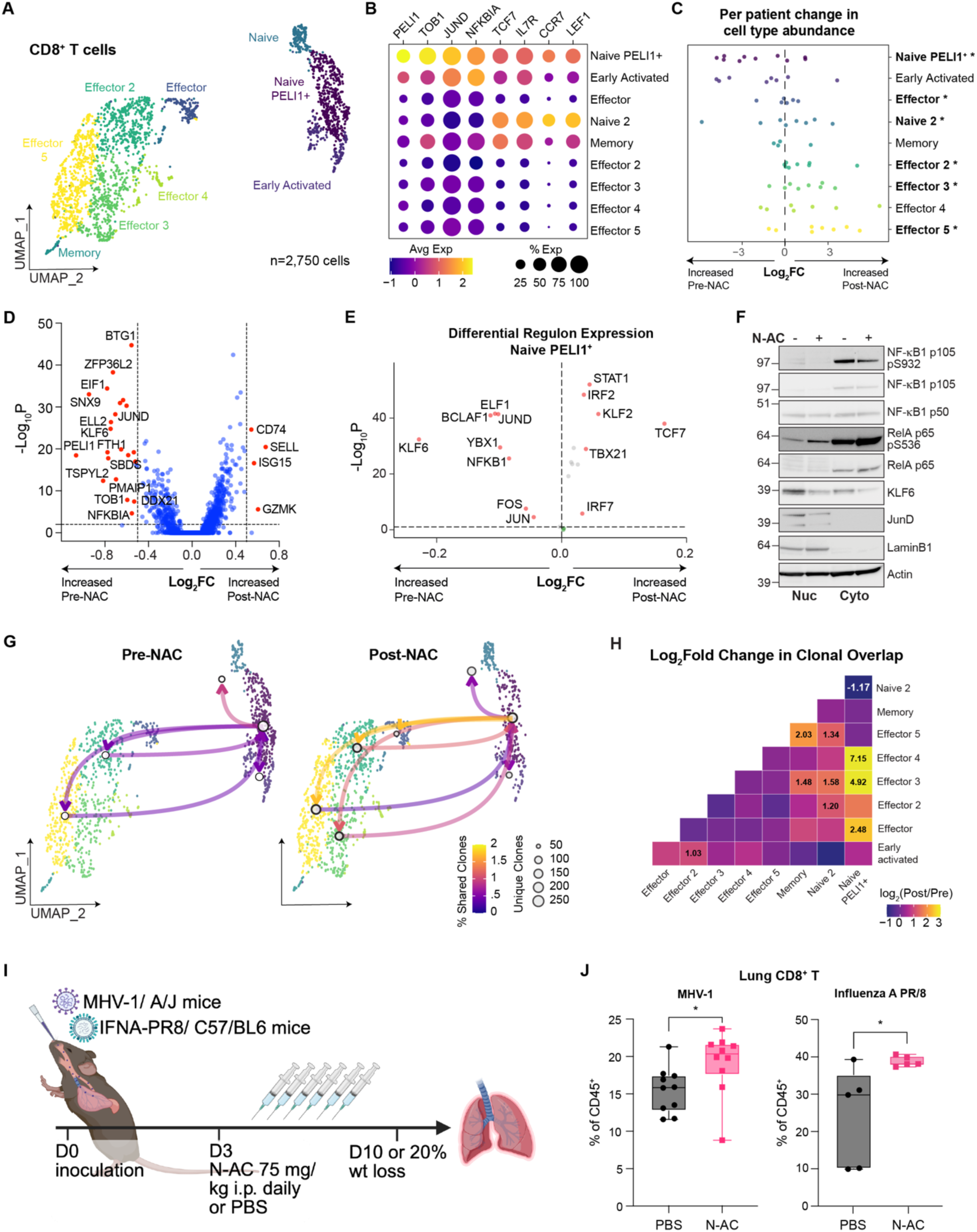
Redox stress limits T cell priming by driving aberrant differentiation of naïve CD8+ T cells. **(A)** UMAP of CD8+ T cells (n = 2,750 cells), based on weighted transcriptomic and gene regulatory profiles inferred by SCENIC. **(B)** Dot plot of scaled expression of genes classically associated with naïve T cells. **(C)** Relative change in cell type abundances per patient in pre-versus on-treatment samples. **(D)** Volcano plot of differentially expressed genes in CD8+ T cells identified by pseudobulk analysis of pre-versus on-treatment samples. **(E)** Volcano plot of differentially expressed transcription factors inferred by SCENIC in the Naïve Peli1+ CD8+ T cells identified by pseudobulk analysis of pre-treatment versus on-treatment samples. **(F)** Nuclear and cytosolic abundance of key differentially expressed transcription factors identified by SCENIC in (E) in OT-I CD8+ T cells activated in the presence or absence of N-AC for 48 hours. **(G)** Network connection of clones shared between Naïve Peli1+ CD8+ T cells and all other CD8+ T cell clusters pre-treatment (left) and on treatment (right). **(H)** Change in pairwise clonal overlap between CD8+ T cell clusters from post-treatment versus pre-treatment samples, calculated using the clonalOverlap function from scRepertoire (method = “morisita”). **(I)** Experimental design used to determine the effect of N-AC on CD8+ T cell responses during *in vivo* MHV-1 or Influenza A PR/8 infection. **(J)** Changes in proportions of CD8+ T cells in lung tissue of MHV-1 (left, N=10 per group) or Influenza A PR/8 (right, N=5 per group) infected mice treated with N-AC or PBS control. *p<.05 **p<.01 ***p<.001 P values determined by unpaired t-test (J). Significant results from scCODA analysis are shown in bold with an asterisk (C).

We next employed gene regulatory network analysis to understand the redox-sensitive transcriptional drivers of this novel “suppressed” T cell state. We found that N-AC treatment led to downregulation of transcription factors with established roles as repressors of T cell proliferation and cytokine production (e.g. JUND, FOXP1, NFKB1) (**Fig. 4E, fig. S4G-H, and Table S16**). Notably, Kruppel-like family factor 6 (KLF6), a quiescence-associated transcription factor that can be induced by oxidative stress (*47*), was the transcription factor whose target genes were most significantly downregulated following N-AC treatment, suggesting that redox stress might limit T cell activation by driving differentiation into a suppressive, naïve-like PELI1+ state. Consistent with this hypothesis, N-AC supplementation during T cell activation reduced nuclear accumulation of KLF6 and JUND, while increasing phosphorylation of the p65 subunit of NF-kB (RelA) (**Fig. 4F**). NF-kB dimer composition instructs transcriptional activity, with p50/p50 homodimers lacking transactivation activity and functioning as transcriptional repressors, while p65/p50 heterodimers promote T-cell activation and cytokine production(*48, 49*), suggesting that N-AC regulates NF-kB signaling by tuning the p65/p50 ratio.

We observed significant expansion of multiple effector T cell subsets, most notably the Effector state, which increased from 10.6% to 20.4%, and the Effector 5 state, which increased from 3.6% to 7.7% (**Fig. 4B, S4D** and **Table S14**). To determine whether reversing redox stress restored the proliferative capacity of PELI1+ naïve T cells, we broadly analyzed T cell clonality prior to and following N-AC treatment using several well-validated TCR diversity metrics(*50*). Treatment was associated with decreases in the Shannon, Inverse-Simpson, Gini-Simpson indices and normalized entropy; all of which correlate negatively with TCR evenness and richness and whose decrease is consistent with both an increase in the number of TCR clones and an expansion of select clones (**Fig. S4I**). We next used scRepertoire to determine whether N-AC promoted expansion and differentiation of specific T cell subsets (*51*). Clonal overlap between naïve, memory, and naïve PELI1+ T cells and effector T cells increased substantially following N-AC treatment, whereas overlap between naïve and naïve PELI1+ T cells decreased, indicating that N-AC treatment promoted T cell expansion and effector differentiation. Cross-referencing of TCRb sequences with TCRb sequences identified as COVID-19 reactive across public databases (*52–55*) revealed that 14% (386/2,750) of identified CD8 TCRs mapped to a COVID-reactive TCRb sequence (**Fig. S4J,K**). Given that majority of COVID-specific TCRs are private to an individual(*52*), this degree of overlap provides confidence that our findings are functionally relevant. Importantly, only 9% (36/386) of these putative COVID-reactive clones also mapped to either EBV or CMV-reactive TCRb sequences. Moreover, nearly all highly expanded (clone size >100) TCR clones within our dataset mapped to TCRs with known COVID-reactive TCRb sequences, consistent with most clonal expansion occurring in COVID-reactive T cells (**Fig. S4J,K**). Together, our data suggests N-AC treatment relieves constraints on activation and enables differentiation into a broader range of effector fates (**Fig. S4L**).

Finally, to orthogonally confirm that redox stress limits T cell activation and effector differentiation in severe respiratory infections, we tested the impact of N-AC administration in mice infected with two distinct respiratory viruses: murine hepatitis virus-1 (MHV-1), a murine coronavirus which reproduces several clinical aspects of COVID-19 infection in humans(*56*), and the PR/8 strain of Influenza A, which causes dose-dependent severe disease and antigen-specific CD8+ T cell immunity in C57/Bl6 mice (*57, 58*). Because these mouse strains are immunocompetent at baseline and increased T cell responses are clearly linked to disease severity in immunocompetent patients(*7, 59*), we restricted our experimental design to determine solely whether N-AC administration was sufficient to increase T cell responses in mice bearing respiratory viral infections. Mice infected with either MHV-1 or Influenza A PR/8 received intraperitoneal N-AC (75 mg/kg/day) or PBS beginning 3 days post-inoculation and were sacrificed 7 days later to determine whether N-AC administration was sufficient to enhance the endogenous CD8^+^ T cell response (**Fig. 4I**). N-AC was sufficient to increase total CD8^+^ as well as activated CD8^+^ (CD8^+^CD44^+^) T cell abundance within the lungs of both MHV-1 infected and Influenza A PR/8 infected mice without evidence of clinically evident immunopathology as demonstrated by the fact that weight loss was not noticeably altered by treatment (**Fig. 4J and S4M-O**).

## DISCUSSION

In this manuscript, we identify cysteine availability as a key driver of adaptive immune dysfunction in cancer patients with severe COVID-19 infection. Intravenous cysteine supplementation reversed metabolic and immunological hallmarks of refractory disease and led to substantial clinical improvement, particularly in patients not yet requiring positive pressure ventilation. Severe viral infections, including COVID-19 remain a substantial cause of morbidity and mortality for several high-risk populations, including older individuals, and patients with comorbid conditions like cancer. These patients have weaker immune responses to vaccination(*60, 61*) and tend to be refractory to therapies aimed at curbing excess inflammation, such as dexamethasone and IL-6 antagonism, underscoring the importance of identifying alternative treatment strategies.

Our findings suggest that morbidity in patients with steroid refractory COVID-19 may be fundamentally driven by blunted adaptive immunity; this is consistent with studies showing that steroid-refractory patients were more likely to have severe disease with higher viral burdens(*62*). Our findings indicate that MHC-II^low^ myeloid cells and CD8+ T cell lymphopenia, which are the strongest predictors of COVID-19 disease severity and treatment response, are driven by redox dysfunction, as they were reversible with N-AC supplementation. However, therapeutic enhancement of CD8+ T cell responses to viral infection should be approached with caution.

Given that overactive T cell responses have also been implicated in COVID-19 morbidity and mortality, particularly in immunocompetent patients, we would suggest that consideration of N-AC or other therapies that enhance anti-viral T cell function are most appropriate in patients with evidence of baseline immune compromise, including those with cancer. This somewhat narrow therapeutic window may explain the mixed results of clinical trials of N-AC in unselected patient populations with COVID-19 (*63, 64*).

Our work suggests that unique redox stress-dependent networks may impair T cell priming through independent effects on both antigen presenting and T cells. We identify the metabolic and oxidative stress response modules, ATF4 and HIF1A, as key drivers of monocyte dysfunction. While neither of these transcription factors directly suppress antigen presentation, both drive expression of PGE2 in myeloid cells, leading to autocrine suppression of MHC expression(*65, 66*). Our observation that PGE2 modulates the threshold for successful T cell activation suggests that PGE2 limits the diversity of T cell responses to cognate antigen, which is clinically significant given that a diverse COVID-19 specific T cell repertoire is predictive of enhanced T cell persistence and improved outcomes(*55, 67–69*). Our data also implicates redox stress in constraining T cell activation through mobilization of stress-associated transcription factors such as KLF6, JUND, and NF-KB1 in CD8 T cells. This transcriptional reprogramming is associated with increased expression of negative regulators of T cell activation, including PELI1 and TOB1, which are typically induced during peripheral tolerance to prevent autoimmunity(*44, 46*). In combination with our mouse studies showing that N-AC increases T cell activation and effector differentiation, these data demonstrate how redox stress can induce tolerogenic signaling that supports quiescence of naïve T cells and anergy of early activated T cells despite high pathogen load.

We acknowledge the potential limitations to this study; the trial was a non-randomized phase II study in which all patients received treatment and therefore the improvement in patients receiving treatment must be interpreted with caution. While the characteristics and treatment history of the pre-trial cohort used for comparison support an interpretation in which N-AC improved clinical outcomes, improved supportive care measures and more prompt administration of standard therapies may also have contributed to improved outcomes. As such, randomized studies will be needed to directly confirm this hypothesis. Furthermore, whether the transcription factors associated with myeloid and T cell dysfunction exhibit redox-dependent DNA binding and gene transactivation requires further exploration. Finally, whether the redox-sensitive T cell dysfunction observed in this patient cohort can be generalized to all patients with cancer or more broadly to all patients with severe disease remains to be determined. Nevertheless, our findings indicate that targeting metabolic immune dysfunction is a tractable strategy in patients with refractory COVID-19.

## MATERIALS AND METHODS

### Study Design

#### Patient cohorts for unbiased proteomic and metabolomic analysis of severe COVID-19

The INCOV cohort included 209 SARS-CoV-2 patients (50% females, aged between 18 and 89 years with an average of 56 years), an expansion on the cohort previously published at acute infection (Su et al., 2020). Potential participants were identified at five hospitals of Swedish Medical Center and affiliated clinics located in the Puget Sound region near Seattle, WA. All enrolled patients provided written in-person informed consent. De-identified proteomic and metabolomic data from matched healthy controls processed using the shared technical pooled control samples to enable batch-correction were previously collected from individuals enrolled in a wellness program (Manor et al., 2018) (Arivale, Seattle, WA). Healthy control samples for single-cell analyses were obtained from Bloodworks Northwest (Seattle, WA). Disease severity was quantified using the WHO Ordinal Scale for Clinical Improvement score (WOS) (World Health Organization, 2020). Clinical data for hospitalized patients were abstracted from deidentified electronic health records (EHR). Clinical lab data were extracted from the nearest time point to each blood draw. Procedures for the INCOV study were approved by the Institutional Review Board (IRB) at Providence St. Joseph Health with IRB study number STUDY2020000175 and the Western Institutional Review Board (WIRB) with IRB study number 20170658. The MSKCC cohort included patients consented to the MSKCC institutional biobanking protocol (IRB #06-107) from whom plasma samples were collected between April 1 and May 31, 2020. Detailed demographic information on both the INCOV and MSKCC patient cohorts are listed in **Table S1**.

### N-Acetylcysteine Phase II Study

#### Enrollment criteria

This was a single institution phase II study evaluating the efficacy of N-AC in hospitalized patients with severe COVID-19 infection. There were two arms to the trial. Arm A consisted of mechanically ventilated patients or patients admitted to the intensive care unit (ICU) (i.e., high concern for respiratory decompensation). Arm B consisted of patients requiring Supplementary oxygen of 2L of more by nasal cannula (to maintain an SpO2 of 95%) but not already admitted to the ICU. Other inclusion criteria required patients to have a documented COVID-19 infection and were ≥18 years of age. For arm A, it was also required that patients have an absolute lymphocyte count (ALC) of >1 cell/µL (patients with lymphoid malignancy were allowed to enroll regardless of ALC). Otherwise, there were no other strict inclusion or exclusion criteria. Patients were allowed to receive any form of concomitant anti-COVID therapies at any time point including steroids, anti-viral therapies, monoclonal antibodies, convalescent plasma, and tocilizumab. The protocol was approved by the Memorial Sloan Kettering internal review board (IRB) and registered on clinicaltrials.gov (NCT04374461).

#### Sample size determination

A Simon optimal design was employed to evaluate the clinical benefit (defined as not requiring ICU or ICU-level care) for each arm of the study. For Arm A (ICU and/or mechanically ventilated patients), it was estimated that the population clinical benefit rate by this definition was 20% and a positive study would be 40%. The parameters of the design were specified as follows: a 20% not promising response rate, a 40% promising response rate, the probability of a type I error (falsely accepting a non-promising therapy) of 0.05, and the probability of a type II error (falsely rejecting a promising therapy) of 0.20. In the first stage of this design, the accrual goal was 13 patients. If at least 4 patients were discharged from the ICU or ICU-based care, then an additional 30 patients would be accrued to the second stage. At the end of the trial, if 13 or more patients were discharged from the ICU or ICU-level care then N-acetylcysteine would be considered worthy of further investigation. This design yielded at least a 0.80 probability of a positive result if the true clinical benefit rate was at least 40% and a 0.95 probability of a negative result if the true clinical benefit rate was 20%. Patients who die in the ICU were considered failures.

For Arm B (non-ICU, non-mechanically ventilated patients), it was estimated that the population clinical benefit rate by this definition was 60% and a positive study would be 80%. The parameters of the design were specified as follows: a 60% not promising response rate, an 80% promising response rate, the probability of a type I error (falsely accepting a non-promising therapy) of 0.05, and the probability of a type II error (falsely rejecting a promising therapy) of 0.20. In the first stage of this design, the accrual goal was 11 patients. If at least 7 patients did not require the ICU or ICU-based care, then an additional 30 patients would be accrued to the second stage. At the end of the trial, if 30 or more patients did not require ICU or ICU-level care then N-acetylcysteine would be considered worthy of further investigation. This design yielded at least a 0.80 probability of a positive result if the true clinical benefit rate is at least 80% and a 0.95 probability of a negative result if the true clinical benefit rate was 60%. Patients who died were considered failures.

#### Intervention

Patients received 6 g/day of IV N-AC as a continuous infusion for up to 21 days. Patients who weighed less than 45 kg had a starting dose reduction of 100 mg/kg over 24 hours for each dose. Criteria for dose reduction or delays are provided in the Supplementary material.

#### Endpoints

Patients received a maximal duration of 21 days of continuous N-AC infusion. Patients meeting predetermined endpoints prior to the maximal 21 days were taken off protocol and deemed either a treatment success or failure. Patients who received the maximal 21 days were determined as a clinical success or failure at the time of their first outcome event. For arm A, successful events included having either been transferred out of the ICU or been extubated, and failures included experiencing either a treatment-related toxicity or death. For arm B, successes included being discharged from the hospital, and failures included an admission to the ICU, being intubated, experiencing a treatment-related toxicity or death. The last patient was enrolled on 4/9/2021 and the trial closed to accrual on 7/19/2021 due to slowed enrollment. Survival records were locked as of 12/1/2021.

#### SARS-CoV-2 RNA testing

SARS-CoV-2 RNA was detected in nasopharyngeal swabs (NPS) or saliva samples using a laboratory-developed test as previously described^5^. Testing was also performed using several commercial assays including the TaqPath™ COVID-19 Combo Kit (Thermo Fisher Scientific, Waltham, MA) targeting the N, S and ORF genes, the cobas® SARS-CoV-2 test (Roche Molecular Diagnostics, Indianapolis, IN) targeting the ORF1 a/b and E gene and the Xpert Xpress SARS-CoV-2 test (Cepheid, Sunnyvale, California) targeting the N and E genes. Samples were reported as positive per manufacturers’ instructions. The cycle threshold (Ct) value, a semi-quantitative estimate of the viral SARS-CoV-2 RNA load that was normalized to internal standards, was retrieved for all gene targets from each instrument record and was confirmed to be internally consistent across commercial assay kits.

### O-link proteomic analysis of plasma

O-link analysis of circulating plasma protein concentrations was performed using a proximity extension assay (PEA) (Olink Proteomics, Uppsala, Sweden) which allows for the simultaneous analysis of 92 protein biomarkers on each panel. Briefly, one microliter of plasma from freshly thawed samples was incubated overnight and allowed to bind with oligonucleotide-labeled antibody pairs to form specific DNA duplexes. This template was then extended and pre-amplified, and the individual protein markers were measured using high-throughput microfluidic real-time PCR (Fluidigm, South San Francisco, CA). The resulting Ct values were normalized and adjusted with a correction factor according to the manufacturer’s instructions to calculate a normalized protein expression value (NPX) in log2 scale. Samples were processed in batches with pooled quality control samples included in each batch. For Fig. 1, the plasma concentrations of proteins were measured using the following Olink Target panels: Cardiometabolic, Cardiovascular II, Cardiovascular III, Cell Regulation, Development, Inflammation, Immune response, Immuno-oncology, Metabolism, Neurology, Neurology Exploratory, Organ Damage, Oncology II, and Oncology III. For Fig. 2, plasma concentrations of proteins were measured using the ProSeek Inflammation and Immune Response panels.

### Metabolomics analysis of plasma

#### Metabolon analysis of plasma

Metabolon measured metabolite levels from plasma samples using ultra high-performance liquid chromatography/tandem accurate mass spectrometry (UHPLC/MS/MS) methods. Metabolon performed all metabolite identification, quantification, and normalization. Pooled quality control samples were included in each batch. To correct for batch effects, metabolite levels were divided by corresponding average values in the pooled quality control within each batch.

#### Targeted metabolomic analysis of plasma samples from NCT04374461

Targeted metabolomic analysis was performed as previously described(*70*). Briefly, plasma isolated from the peripheral blood of patients on NCT04374461 was frozen and stored at -80C. Samples were subsequently thawed, and metabolites were extracted from 100 uL of plasma by combining with 900 uL of ice-cold 80% methanol. After overnight incubation at -80**°**C, samples were centrifuged at 21,000*g* for 20 minutes and 900 uL of supernatant was decanted to a fresh Eppendorf tube to remove protein. Extracts were dried in an evaporator (Genevac EZ-2 Elite) and were subsequently resuspended in 60 μl of 60% acetonitrile in water for hydrophilic interaction liquid chromatography (HILIC). Samples were vortexed, incubated on ice for 20 min and clarified by centrifugation at 20,000g for 20 min at 4°C. HILIC LC–MS analysis was performed on a 6545 Q-TOF mass spectrometer (Agilent Technologies) in both positive and negative ionization modes using columns, buffers and LC-MS parameters as described previously (*70*). Peak identification and integration were done based on exact mass and retention time match to commercial standards. Targeted data analysis was performed using Skyline v.25.1. For profiling experiments, peak areas of 13C15N-labeled amino acid internal standards were analyzed to confirm <10% of inter-sample variability.

### Consensus NMF Topic Modeling

To identify latent modules within plasma metabolomic and proteomic datasets, we applied consensus non-negative matrix factorization (cNMF) using the sklearn.decomposition.NMF implementation in Python. For quality control, we filtered out features with <75% data coverage within any experimental group. The remaining missing values were imputed using k-nearest neighbors (KNN). Data were scaled using MinMaxScaler to ensure non-negativity required for NMF. We evaluated a range of components k (k = 2-10) across 20 independent NMF runs per k using nonnegative double singular value decomposition (NNDSVD) initialization (init=”nndsvd”) and allowing up to 1000 iterations. To account for label switching, sample loading matrices (W) were aligned across runs using pairwise correlation and the Hungarian algorithm (scipy.omtimize.linear_sum_assignment). We chose k=4 and k=7, as the optimal number of components for O-link and Metabolomics NMF, respectively, based on the elbow of the reconstruction error curve and high stability score. The stability score was calculated as the standard deviation of the aligned distance matrices over 20 runs. The final consensus W (module score per sample) and H (feature weights per module) matrices were calculated by averaging across the 20 aligned NMF runs at our chosen k.

### Animal models

All animal experiments were performed according to Memorial Sloan Kettering Cancer Center Institutional Animal Care and Use Committee (IACUC) guidelines (Protocol Number 20-10012). Mice were used at between 8 and 12 weeks of age unless otherwise indicated. Age and sex matched mice were used in each experiment. The following mice were housed and bred under specific pathogen-free conditions at Memorial Sloan Kettering Cancer Center (MSKCC) barrier facility: C57BL/6J (Strain # 000664), C57BL/6-Tg(TcraTcrb)1100Mjb/J (OT-1) (Strain # 003831), and A/J (Strain # :000646) mice were purchased from The Jackson Laboratory.

#### PR8 Influenza model

PR8 viral stock was kindly provided by the Jayanta Chaudhuri lab. C57BL/6J (Strain # 000664) (8-12wks old) were inoculated intranasally with 50 TCID50 PR8 diluted in PBS to 25 uL. At 3d later, mice were intraperitoneally injected daily with N-Acetylcysteine (75mg/kg) or PBS alone. Mice were monitored daily and killed for signs of morbidity.

#### MHV Coronavirus model

MHV viral stock was kindly provided by the Joe Sun lab. A/J (Strain # :000646) (8-12 wks old) were inoculated intranasally with 1×10^5^ PFU MHV diluted in PBS to 25 uL. At 3d later, mice were intraperitoneally injected daily with N-AC (75 mg/kg) or PBS alone. Mice were monitored daily and killed for signs of morbidity.

### Human Single Cell Transcriptomics

#### Gene Regulatory Network Analysis

SCENIC was applied to filtered and normalized RNA count matrices from subset populations (pySCENIC). Motif databases and transcription factor lists were downloaded from resources.artslab.org (v9) and regulon prediction was done using auc_threshold of 0.05. Output matrices were combined into subset Seurat objects. PCA was performed using Seurat’s Run PCA function and to mitigate effect of lane-specific and sample-specific covariates/batch effects, Harmony v.1.0.0 RunHarmony was used to correct embedding. Elbow plots and jack straw plots were manually inspected to determine the number of principal components to use downstream and a global UMAP was constructed using Seurat’s weighted nearest-neighbor workflow upon corrected RNA and SCENIC data.

#### CellphoneDB Interaction Pair Analysis

CellphoneDBv5 was applied to infer ligand-receptor interactions driven by statistically significant upregulated genes. Differentially expressed genes between timepoints were calculated per cell type using Seurat FindMarkers and filtered for adjusted p-value (FDR) <0.05, log 2 fold change >0.2, and expressed in a minimum of 10% of cells per cluster. CellphoneDB requires positively upregulated genes, so we ran in parallel 2 implementations of CellphoneDB with DEGs significantly upregulated in Timepoint A compared to Timepoint B and vice versa.

#### Gene Signature Analysis

Treatment induced transcriptional changes were calculated using Seurat’s implementation of the Wilcoxon rank-sum test (Find Markers). Fgsea v.1.24.0 and escape v1.8.0 were used to evaluate-functional module level expression changes of various gene sets taken from the Molecular Signatures Database (MSigDB).

## List of Supplementary Materials

Supplementary Materials and Methods

Fig. S1 to S4

Supplementary Tables 1-16

## Acknowledgments

We thank members of the Vardhana laboratory for productive discussion. Influenza PR/8 viral stock was kindly provided by the Jayanta Chaudhuri lab. MHV-1 viral stock was kindly provided by the Joseph Sun lab. J.D.W. acknowledges support from the Ludwig Collaborative and Swim Across America Laboratory and the Parker Institute for Cancer Immunotherapy.

## Author contributions

Conceptualization: TRM, WTJ, SAV Methodology: TRM, JR, WTJ, MB

Investigation: TRM, LM, JZ, YHL, WTJ, LM, SG, JN, EC, KKH, GS, JJB, NEB, JDW, JH

Visualization: TRM, JH, AA, MB, JC, SAV

Statistics: HLK, KP

Funding acquisition: TRM, SAV

Supervision: SAV, BG

Writing – original draft: TRM, SAV

Writing – reviewing & editing: TRM, SAV

## Data and materials availability

Sequencing data will be available on dbGaP at the time of publication. All clinical data generated during and/or analyzed during the current study are available from the corresponding author on reasonable request. All other data are available in the main text or the supplementary materials.

## References and Notes

1. J. L. Wong, S. E. Evans, Bacterial Pneumonia in Patients with Cancer: Novel Risk Factors and Management. Clin Chest Med 38, 263–277 (2017).

2. R. J. P. Jose et al., Cancer patients with community-acquired pneumonia treated in intensive care have poorer outcomes associated with increased illness severity and septic shock at admission to intensive care: a retrospective cohort study. Pneumonia (Nathan) 6, 77–82 (2015).

3. S. Tanaka et al., Increased risk of death from pneumonia among cancer survivors: A propensity score-matched cohort analysis. Cancer Med 12, 6689–6699 (2023).

4. N. M. Kuderer et al., Clinical impact of COVID-19 on patients with cancer (CCC19): a cohort study. Lancet 395, 1907–1918 (2020).

5. E. V. Robilotti et al., Determinants of COVID-19 disease severity in patients with cancer. Nat Med 26, 1218–1223 (2020).

6. M. Merad, C. A. Blish, F. Sallusto, A. Iwasaki, The immunology and immunopathology of COVID-19. Science 375, 1122–1127 (2022).

7. R. C. Group et al., Dexamethasone in Hospitalized Patients with Covid-19. N Engl J Med 384, 693–704 (2021).

8. T. F. Aiello et al., Dexamethasone treatment for COVID-19 is related to increased mortality in hematologic malignancy patients: results from the EPICOVIDEHA registry. Haematologica 109, 2693–2700 (2024).

9. D. R. Rivera et al., Utilization of COVID-19 Treatments and Clinical Outcomes among Patients with Cancer: A COVID-19 and Cancer Consortium (CCC19) Cohort Study. Cancer Discov 10, 1514–1527 (2020).

10. E. M. Bange et al., CD8(+) T cells contribute to survival in patients with COVID-19 and hematologic cancer. Nat Med 27, 1280–1289 (2021).

11. S. DeWolf et al., SARS-CoV-2 in immunocompromised individuals. Immunity, (2022).

12. C. Y. Lee et al., Prolonged SARS-CoV-2 Infection in Patients with Lymphoid Malignancies. Cancer Discov 12, 62–73 (2022).

13. O. Lyudovyk et al., Impaired humoral immunity is associated with prolonged COVID-19 despite robust CD8 T cell responses. Cancer Cell 40, 738–753 e735 (2022).

14. J. M. Amrute et al., Cell specific peripheral immune responses predict survival in critical COVID-19 patients. Nat Commun 13, 882 (2022).

15. A. Unterman et al., Single-cell multi-omics reveals dyssynchrony of the innate and adaptive immune system in progressive COVID-19. Nat Commun 13, 440 (2022).

16. A. Palazon, A. W. Goldrath, V. Nizet, R. S. Johnson, HIF transcription factors, inflammation, and immunity. Immunity 41, 518–528 (2014).

17. Y. Su et al., Multi-Omics Resolves a Sharp Disease-State Shift between Mild and Moderate COVID-19. Cell 183, 1479–1495 e1420 (2020).

18. Y. Su et al., Multiple early factors anticipate post-acute COVID-19 sequelae. Cell 185, 881–895 e820 (2022).

19. D. D. Lee, H. S. Seung, Learning the parts of objects by non-negative matrix factorization. Nature 401, 788–791 (1999).

20. M. Merad, J. C. Martin, Pathological inflammation in patients with COVID-19: a key role for monocytes and macrophages. Nat Rev Immunol 20, 355–362 (2020).

21. S. S. Tate, E. M. Grau, A. Meister, Conversion of glutathione to glutathione disulfide by cell membrane-bound oxidase activity. Proc Natl Acad Sci U S A 76, 2715–2719 (1979).

22. M. Emdin et al., Prognostic value of serum gamma-glutamyl transferase activity after myocardial infarction. Eur Heart J 22, 1802–1807 (2001).

23. A. Zorlu et al., Increased gamma-glutamyl transferase levels predict early mortality in patients with acute pulmonary embolism. Am J Emerg Med 30, 908–915 (2012).

24. J. W. Lee et al., Integrated analysis of plasma and single immune cells uncovers metabolic changes in individuals with COVID-19. Nat Biotechnol 40, 110–120 (2022).

25. P. Jouandin et al., Lysosomal cystine mobilization shapes the response of TORC1 and tissue growth to fasting. Science 375, eabc4203 (2022).

26. A. Varghese et al., Unravelling cysteine-deficiency-associated rapid weight loss. Nature, (2025).

27. S. A. Vardhana et al., Impaired mitochondrial oxidative phosphorylation limits the self-renewal of T cells exposed to persistent antigen. Nat Immunol 21, 1022–1033 (2020).

28. Q. Lu et al., SARS-CoV-2 exacerbates proinflammatory responses in myeloid cells through C-type lectin receptors and Tweety family member 2. Immunity 54, 1304–1319 e1309 (2021).

29. L. Tang et al., Liver sinusoidal endothelial cell lectin, LSECtin, negatively regulates hepatic T-cell immune response. Gastroenterology 137, 1498–1508 e1491–1495 (2009).

30. M. Buttner, J. Ostner, C. L. Muller, F. J. Theis, B. Schubert, scCODA is a Bayesian model for compositional single-cell data analysis. Nat Commun 12, 6876 (2021).

31. E. Dann, N. C. Henderson, S. A. Teichmann, M. D. Morgan, J. C. Marioni, Differential abundance testing on single-cell data using k-nearest neighbor graphs. Nat Biotechnol 40, 245– 253 (2022).

32. R. Knoll et al., The life-saving benefit of dexamethasone in severe COVID-19 is linked to a reversal of monocyte dysregulation. Cell 187, 4318–4335 e4320 (2024).

33. S. E. Holton et al., Mediators of monocyte chemotaxis and matrix remodeling are associated with mortality and pulmonary fibroproliferation in patients with severe COVID-19. PLoS One 19, e0285638 (2024).

34. S. Aibar et al., SCENIC: single-cell regulatory network inference and clustering. Nat Methods 14, 1083–1086 (2017).

35. B. Van de Sande et al., A scalable SCENIC workflow for single-cell gene regulatory network analysis. Nat Protoc 15, 2247–2276 (2020).

36. K. Troule et al., CellPhoneDB v5: inferring cell-cell communication from single-cell multiomics data. Nat Protoc, (2025).

37. M. Ricke-Hoch et al., Impaired immune response mediated by prostaglandin E2 promotes severe COVID-19 disease. PLoS One 16, e0255335 (2021).

38. S. B. Lacher et al., PGE(2) limits effector expansion of tumour-infiltrating stem-like CD8(+) T cells. Nature 629, 417–425 (2024).

39. M. Morotti et al., PGE(2) inhibits TIL expansion by disrupting IL-2 signalling and mitochondrial function. Nature 629, 426–434 (2024).

40. D. Zehn, S. Y. Lee, M. J. Bevan, Complete but curtailed T-cell response to very low-affinity antigen. Nature 458, 211–214 (2009).

41. M. R. Jenkins, A. Tsun, J. C. Stinchcombe, G. M. Griffiths, The strength of T cell receptor signal controls the polarization of cytotoxic machinery to the immunological synapse. Immunity 31, 621–631 (2009).

42. D. Wu et al., Structural assessment of HLA-A2-restricted SARS-CoV-2 spike epitopes recognized by public and private T-cell receptors. Nat Commun 13, 19 (2022).

43. X. Zhang et al., Viral and host factors related to the clinical outcome of COVID-19. Nature 583, 437–440 (2020).

44. M. Chang et al., The ubiquitin ligase Peli1 negatively regulates T cell activation and prevents autoimmunity. Nat Immunol 12, 1002–1009 (2011).

45. A. Meixner, F. Karreth, L. Kenner, E. F. Wagner, JunD regulates lymphocyte proliferation and T helper cell cytokine expression. EMBO J 23, 1325–1335 (2004).

46. D. Tzachanis et al., Tob is a negative regulator of activation that is expressed in anergic and quiescent T cells. Nat Immunol 2, 1174–1182 (2001).

47. R. Urtasun, F. J. Cubero, N. Nieto, Oxidative stress modulates KLF6Full and its splice variants. Alcohol Clin Exp Res 36, 1851–1862 (2012).

48. S. Grundstrom, P. Anderson, P. Scheipers, A. Sundstedt, Bcl-3 and NFkappaB p50-p50 homodimers act as transcriptional repressors in tolerant CD4+ T cells. J Biol Chem 279, 8460– 8468 (2004).

49. J. H. Lai et al., RelA is a potent transcriptional activator of the CD28 response element within the interleukin 2 promoter. Mol Cell Biol 15, 4260–4271 (1995).

50. J. Mika et al., A comprehensive evaluation of diversity measures for TCR repertoire profiling. BMC Biol 23, 133 (2025).

51. N. Borcherding, N. L. Bormann, G. Kraus, scRepertoire: An R-based toolkit for single-cell immune receptor analysis. F1000Res 9, 47 (2020).

52. S. Nolan et al., A large-scale database of T-cell receptor beta sequences and binding associations from natural and synthetic exposure to SARS-CoV-2. Front Immunol 16, 1488851 (2025).

53. M. Goncharov et al., VDJdb in the pandemic era: a compendium of T cell receptors specific for SARS-CoV-2. Nat Methods 19, 1017–1019 (2022).

54. M. V. Pogorelyy et al., Resolving SARS-CoV-2 CD4(+) T cell specificity via reverse epitope discovery. Cell Rep Med 3, 100697 (2022).

55. A. A. Minervina et al., SARS-CoV-2 antigen exposure history shapes phenotypes and specificity of memory CD8(+) T cells. Nat Immunol 23, 781–790 (2022).

56. A. D. Masciarella, et al., A Mouse Model of MHV-1 Virus Infection for Study of Acute and Long COVID Infection. Curr Protoc 3, e896 (2023).

57. G. T. Belz, P. G. Stevenson, P. C. Doherty, Contemporary analysis of MHC-related immunodominance hierarchies in the CD8+ T cell response to influenza A viruses. J Immunol 165, 2404–2409 (2000).

58. K. J. Flynn et al., Virus-specific CD8+ T cells in primary and secondary influenza pneumonia. Immunity 8, 683–691 (1998).

59. D. Mathew et al., Deep immune profiling of COVID-19 patients reveals distinct immunotypes with therapeutic implications. Science 369, (2020).

60. A. Fendler et al., COVID-19 vaccines in patients with cancer: immunogenicity, efficacy and safety. Nat Rev Clin Oncol 19, 385–401 (2022).

61. L. Monin et al., Safety and immunogenicity of one versus two doses of the COVID-19 vaccine BNT162b2 for patients with cancer: interim analysis of a prospective observational study. Lancet Oncol 22, 765–778 (2021).

62. R. Arcani et al., Factors associated with dexamethasone efficacy in COVID-19. A retrospective investigative cohort study. J Med Virol 94, 3169–3175 (2022).

63. C. S. Kow, D. S. Ramachandram, S. S. Hasan, K. Thiruchelvam, Effect of N-Acetylcysteine on mortality in COVID-19 patients: A systematic review and meta-analysis of randomized controlled trials. Inflammopharmacology 33, 4871–4877 (2025).

64. T. H. Liu et al., Clinical efficacy of N-acetylcysteine for COVID-19: A systematic review and meta-analysis of randomized controlled trials. Heliyon 10, e25179 (2024).

65. M. R. Jeong, J. W. Hwang, M. Choi, S. H. Seok, MHC class II(+) macrophage differentiation is impaired in metastasized lungs via PGE(2) receptor EP2. Cell Rep 44, 115574 (2025).

66. X. H. Liu et al., Prostaglandin E2 induces hypoxia-inducible factor-1alpha stabilization and nuclear localization in a human prostate cancer cell line. J Biol Chem 277, 50081–50086 (2002).

67. K. V. Zornikova et al., Clonal diversity predicts persistence of SARS-CoV-2 epitope-specific T-cell response. Commun Biol 5, 1351 (2022).

68. K. I. Wagner et al., Recruitment of highly cytotoxic CD8(+) T cell receptors in mild SARS-CoV-2 infection. Cell Rep 38, 110214 (2022).

69. A. S. Shomuradova et al., SARS-CoV-2 Epitopes Are Recognized by a Public and Diverse Repertoire of Human T Cell Receptors. Immunity 53, 1245–1257 e1245 (2020).

70. S. Schworer et al., Proline biosynthesis is a vent for TGFbeta-induced mitochondrial redox stress. EMBO J 39, e103334 (2020).

