## Supplementary Materials for "Cysteine supplementation reverses immune dysfunction in cancer patients with severe COVID-19"

### Supplementary Materials and Methods

#### Cell Culture

##### *Cell Isolation for PGE2-Dependent T cell Priming Assay*

For dendritic cell preparation, spleens from 8-10wk C57BL/6 mice were minced and digested in RPMI containing Liberase (Roche, 2.5mg/mL, 1:50) and DNase I (Sigma, 200x) for 30 min at 37°C. In parallel, spleens from 8-10wk OT-1 mice were mechanically dissociated through a 40µm strainer and red blood cells were lysed using ACK lysis buffer. Lysis was quenched with a 10X volume of PBS and centrifuged. OT-I CD8<sup>+</sup> T cells and dendritic cells (DCs) were isolated via negative magnetic selection kits (Dynabeads™ Untouched Mouse T cells Kit, and Dynabeads™ Mouse DC Enrichment Kit, Thermo Fisher), following the manufacturer's instructions.

##### *T Cell-DC coculture*

DCs were resuspended in complete RPMI (RPMI-1640 medium supplemented with 10% fetal bovine serum (FBS), 2nM L-glutamine, and penicillin-streptomycin) and incubated for 1 hour at 37°C with gentle rotation in the presence of titrated concentrations of (1, 10, and 100 nM) of SIINFEKL (N4) and SIIVFEKL (V4) peptides. Cells were washed and resuspended at 1x10<sup>6</sup> cells/mL in complete RPMI. In parallel, OT-I T cells were labeled with CellTrace Violet. OT-I T cells were resuspended at 1x10<sup>6</sup> cells/mL and mixed with Cell Trace Violet (5µM) in pre-warmed PBS. Cells were incubated for 20 min at 37°C and then washed with 5X volume of complete RPMI. CTV-labeled cells were centrifuged and resuspended at 2x10<sup>5</sup> cells/mL in T cell media (complete RPMI supplemented with 50 µM 2-Mercaptoethanol, and 10 ng/mL murine IL-2 (Peprotech). In a 96-well polypropylene deep-well plate (Corning), 5x10<sup>3</sup> peptide-pulsed DCs (5 µL) were combined with 1x10<sup>5</sup> OT-I T cells (500 µL) per replicate. Then, cells were treated with 1, 10, or 100 ng/mL Prostaglandin E2 (Sigma) or DMSO alone. Cells were incubated for 4 days at 37°C.

##### *PMA/Ionomycin Restimulation*

For cytokine analysis, cells were stimulated with 50 ng/mL phorbol 12-myristate 13-Acetate (PMA) and 500 ng/mL ionomycin for 1 hour at 37°C, and then treated with Brefeldin A (1:1000) for 3 hours at 37°C.

##### *Immunophenotyping*

Cells were labeled with viability dye (Ghost Dye 510, Tonbo #13-0870-T500) for 10 minutes at room temperature, then stained with surface markers for 20 minutes at room temperature. For intracellular staining, cells were fixed for 30 min at room temperature with eBioscience FoxP3 Fixation/Permeabilization Kit, and then stained with intracellular markers overnight at 4°C. Next, cells were fixed with 4% formaldehyde in PBS for 10 minutes at room temperature, washed, and resuspended in FACs buffer (PBS with 2% FBS). Data acquisition was performed using a Cytex Aurora spectral flow cytometer, and data were processed using FlowJo v10.10.

##### *Primary mouse T cell isolation and activation*

Primary mouse OT-1 T cells were isolated by negative magnetic isolation as described above. Isolated cells were resuspended at 1.5-2 x 10<sup>6</sup> T cells/mL in T cell media (RPMI-1640 medium supplemented with 10% FBS, 2 mM L-glutamine, penicillin-streptomycin, 50 µM 2-

Mercaptoethanol) with 10 ng/mL murine IL-2 and activated with plate-bound anti-CD3 (2C11, 3 µg/mL) and anti-CD28 (37.51, 1 µg/mL) for 48 hours. N-AC (Sigma) was resuspended in water and used at 10 mM.

##### *Primary Human T cell isolation and activation*

PBMCs were isolated from whole blood (New York Blood Center) by Lymphoprep density gradient-based centrifugation. Primary human T cells were isolated from PBMCs using Dynabeads Untouched Human T cell kit according to manufacturer's instructions. T cell were resuspended in T cell media (RPMI-1640 medium supplemented with 10% FBS, 2 mM L-glutamine, penicillin-streptomycin, 50 µM 2-Mercaptoethanol) with human IL-7 10 ng/mL (PeproTech) and human IL-15 10 ng/mL (PeproTech) and activated with plate-bound anti-CD3 (OKT3, 5 µg/mL) and anti-CD28 (28.2, 2 µg/mL) for 48 hours. For hypoxia experiments, cells were cultured in a hypoxia chamber (Coy) at 0.5% oxygen. N-AC (Sigma) was resuspended in water and used at 10 mM.

##### *Immunoblotting*

Total Protein lysates were extracted in RIPA buffer (Thermo Scientific). Nuclear and cytoplasmic protein lysates were extracted using NE-PER Nuclear and Cytoplasmic Extraction Reagents (Thermo Scientific). Lysates were quantified by BCA (Thermo Fisher), separated by SDS-PAGE and transferred to nitrocellulose (BioRad). Membranes were blocked in 5% milk in Tris (pH)-buffered saline with 0.1% Tween-20 (TBST) and incubated at 4°C with primary antibodies overnight. Membranes were washed with TBST, incubated at room temperature with horseradish peroxidase-conjugated secondary antibodies (Cytiva, GENA934 or Cell Signaling Technology, 7076 at 1:7500) for 1 hour, incubated with enhanced chemiluminescence substrate (Kindle Bioscience or Thermo Scientific), and imaged using a ChemiDoc Touch Imaging System (BioRad). Antibodies used (1:1000, unless otherwise noted) were NFκB1, p105, p50 Polyclonal antibody (Proteintech, 14220-1-AP), Phospho-NF-κB p105 (Ser932) (18E6) (CST, 4806S), NFκB p65 (CST, 8242), Phospho-NF-κB p65 (Ser536) (CST, 3033T), JunD (CST, 5000S), KLF6 (Santa Cruz, sc-365633), Pellino-1 (CST, 31474), Beta-Actin (Sigma, A2228, 1:5000), and LaminB1 (Proteintech, 12987-1-AP, 1:5000).

#### **Human Single Cell Transcriptomics**

##### *Sample Processing*

Frozen peripheral blood mononuclear cells from paired pre-treatment (day 0) and on-treatment (day 7) samples from 13 patients, were thawed and stained with viability dye followed by surface staining with CD45 (Biolegend), TotalSeq-C Human Universal Cocktail (Biolegend) and TotalSeqC human hashtag antibodies. Live CD45+ PBMCs were flow sorted and encapsulated using the Chromium Single Cell Immune Profiling v2 (10X Genomics) and sequenced on NovaSeq system (Illumina).

##### *Data preprocessing, initial processing, and batch correction*

Raw FASTQ data was processed using Cellranger v6.0.2 count workflow to generate gene expression count matrices. TCR data was processed using Cellranger's vdj pipeline to generate cell-clonotype annotations. We used Scrublet for initial doublet detection and removal. Processed RNA and ADT matrixes were combined into a single Seurat v4.0.0 object, and TCR clonotype information was incorporated using scRepertoire combineExpression function. Then, we demultiplexed HTO data with HTODemux and removed doublets and negative cells. We further filtered cells to a minimum feature count of 200, maximum feature count of 3500, and maximum mitochondrial of 5% mitochondrial reads, leaving 20737 cells. RNA and ADT data were normalized using Seurat's NormalizeData function with 'LogNormalize' and 'CLR' methods.

PCA was performed (using Seurat's *RunPCA* function) and to mitigate the effect of lane-specific and sample-specific covariates/batch effects, Harmony v1.0.0 *RunHarmony* function was used to correct embeddings. Elbow plots and jack straw plots were manually inspected to determine the number of principal components to use downstream and a global UMAP was constructed using Seurat's weighted nearest-neighbor workflow upon corrected RNA and ADT data. Using clustree v.0.5.0 produced tree diagrams for various resolution input values, cluster stability was evaluated to avoid over-clustering and optimize the number of communities selected. We removed platelet and red blood cell clusters, leaving 20494 cells.

##### *Population subsetting*

Subsetting was performed by selecting cell clusters from the main Seurat object based on canonical marker expression by RNA and ADT, with concomitant scGate v1.2.0 gating models.

##### *Cell Type Abundance*

To detect statistically credible compositional changes of cell types from our single-cell transcriptomics data we used scCODA(28). The model was fit with timepoint (pre-treatment and on-treatment) and patient as covariates, and platelets as a reference cell type at an FDR level of 0.2. For visualization of changes in cell-type proportions per patient we calculated log<sub>2</sub> fold changes in the proportion of each cell cluster using a generalized linear model (GLM)-based framework. To account for sample-specific differences in sequencing depth, we calculated number of cells per cluster over total cells for each sample (26, 13 patients x 2 timepoints). For each sample, clusters with fewer than 2 cells were excluded to avoid unstable estimates. A small pseudocount ($1 \times 10^{-5}$) was added to minimize technical noise and compositional artifacts. We modeled the proportion of each cluster as the response variable in a GLM with a quasibinomial family and timepoint as the predictor. Log2fc for each patient per cluster is plotted. Clusters were ordered by mean log2fc for visualization on beeswarm plots.

A

### O-link Proteomics

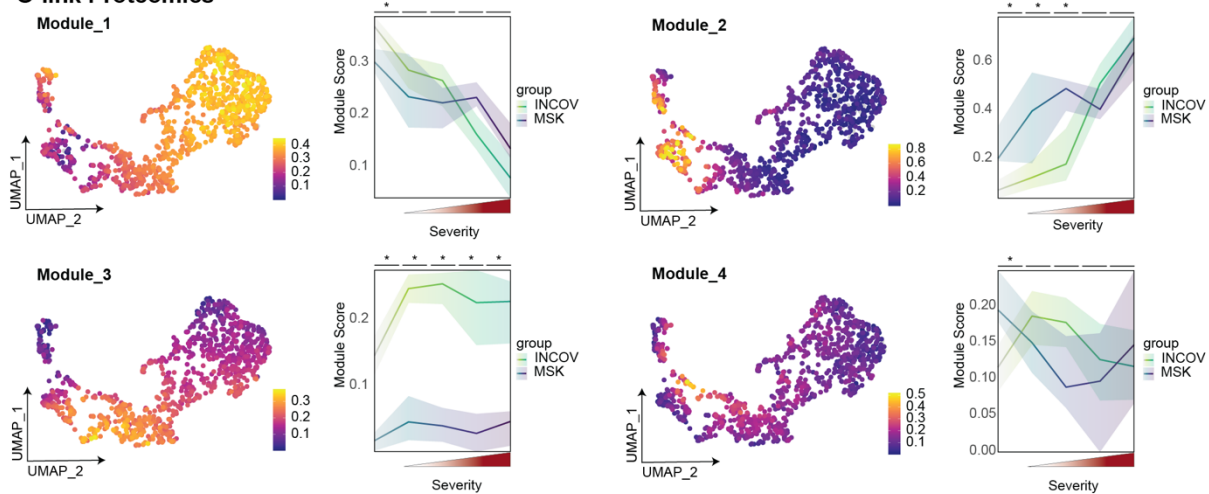

B

### Metabolomics

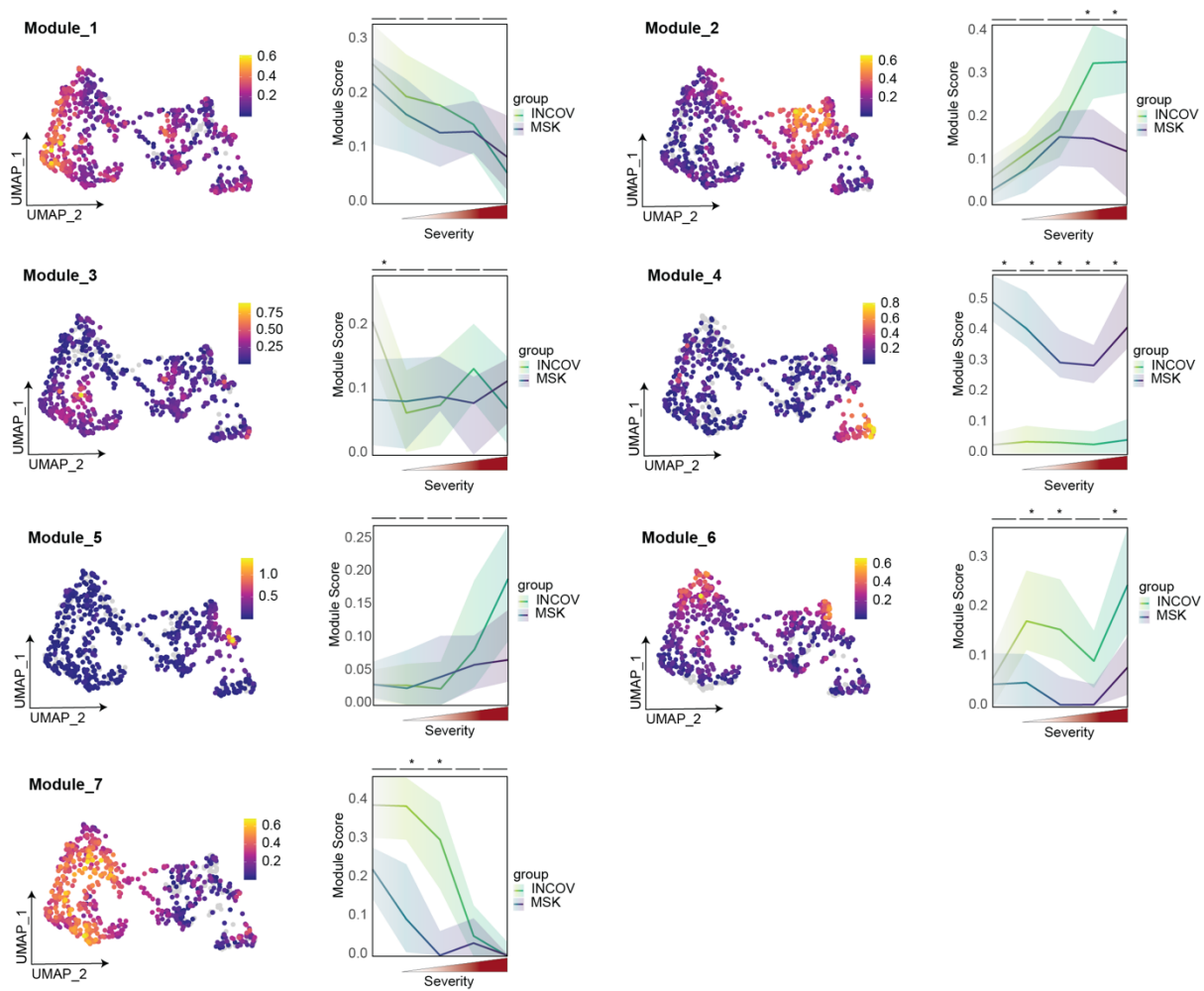

**Supplementary Figure 1. Consensus non-negative matrix factorization identifies COVID-19 severity-associated immune and metabolic signatures in patients with and without cancer. (A)** Expression of Proteomics module scores calculated by consensus NMF per patient overlayed on UMAP projections and corresponding line graphs in which average is bolded, and 25th to 75th percentile highlighted. **(B)** Expression of Metabolomics module scores calculated by consensus NMF per patient overlayed on UMAP projection and corresponding line graphs in which average is bolded, and 25th to 75th percentile highlighted.

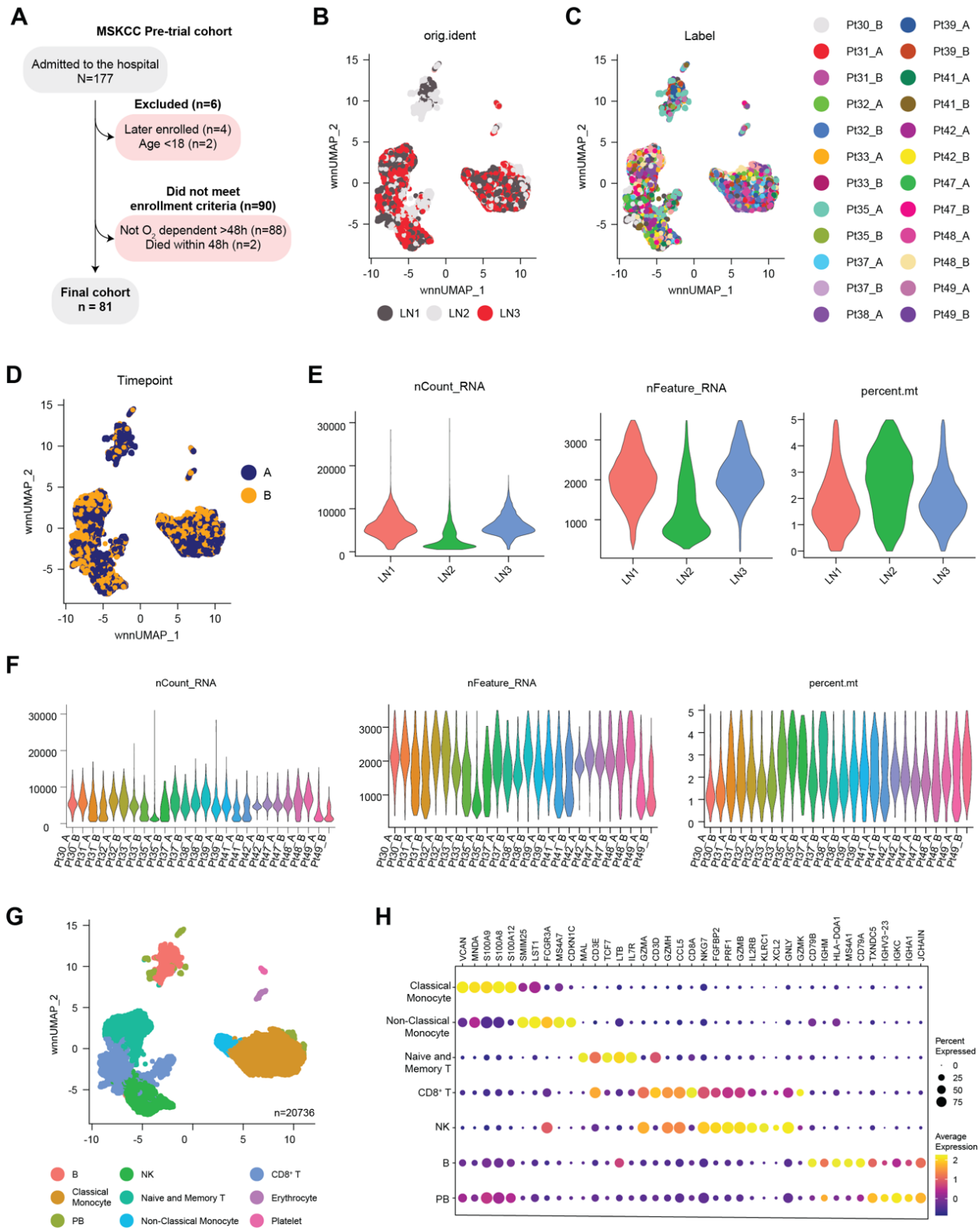

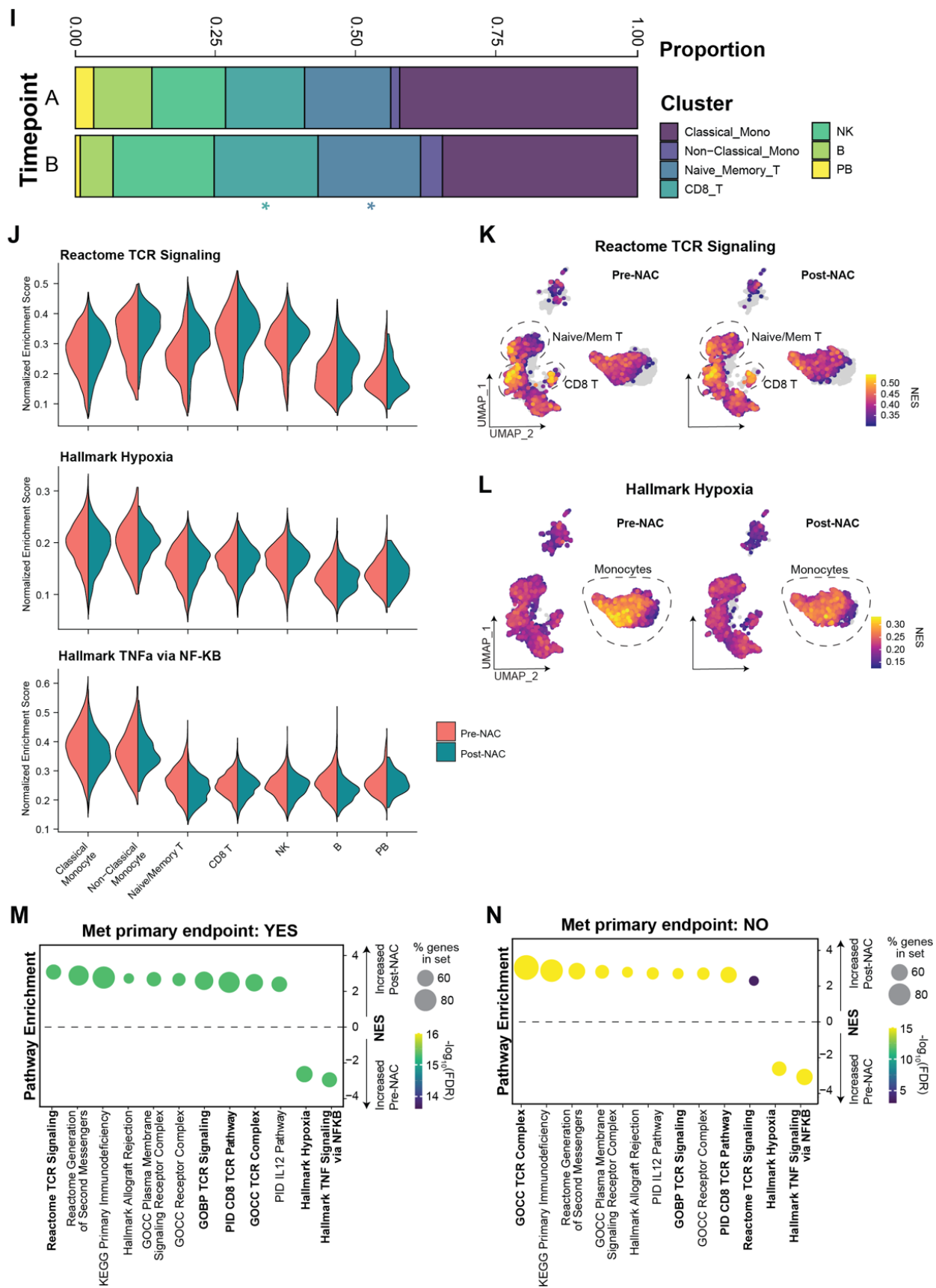

**Supplementary Figure 2. N-Acetylcysteine has differential effects on hypoxia, cytokine, and TCR signaling across immune cell lineages.** (A) Flow chart demonstrating assembly of pre-trial cohort for comparisons in Figure 2. (B-D) UMAP projection of integrated single cell data, labeled by sequencing lane (B), patient sample (C), and timepoint (D). Integration was performed using Harmony. (E,F) Violin plots of single-cell unique molecular identifier counts (nCount\_RNA), detected features (nFeature\_RNA), and mitochondrial transcript percentage across sequencing lanes (E) or individual patient samples (F). (G) U-MAP projection of single cell data labeled by broad cell lineage, including platelets and erythrocytes (n=20,736 cells). (H) Dot plot of expression of marker genes per cluster of single-cell transcriptomic data. (I) Stacked bar plot of cell type proportion changes averaged across samples stratified by Pre-NAC (A) or Post-NAC (B). Differences that are significantly different between timepoints by scCODA are marked with an asterisk. (J) Violin plot of single-cell Reactome TCR Signaling, Hallmark Hypoxia, Hallmark TNFa Signaling via NF-KB, and pathway normalized enrichment scores (Enrichr) split by timepoint within each cluster. (K,L) Expression of Reactome TCR Signaling (K) and Hallmark Hypoxia (L) pathway normalized enrichment scores (Enrichr) in immune cells pre- and on-treatment PBMC samples. (M,N) Enrichment of preranked genesets following initiation of N-AC treatment in patients who did (M) and did not (N) meet the primary endpoint.

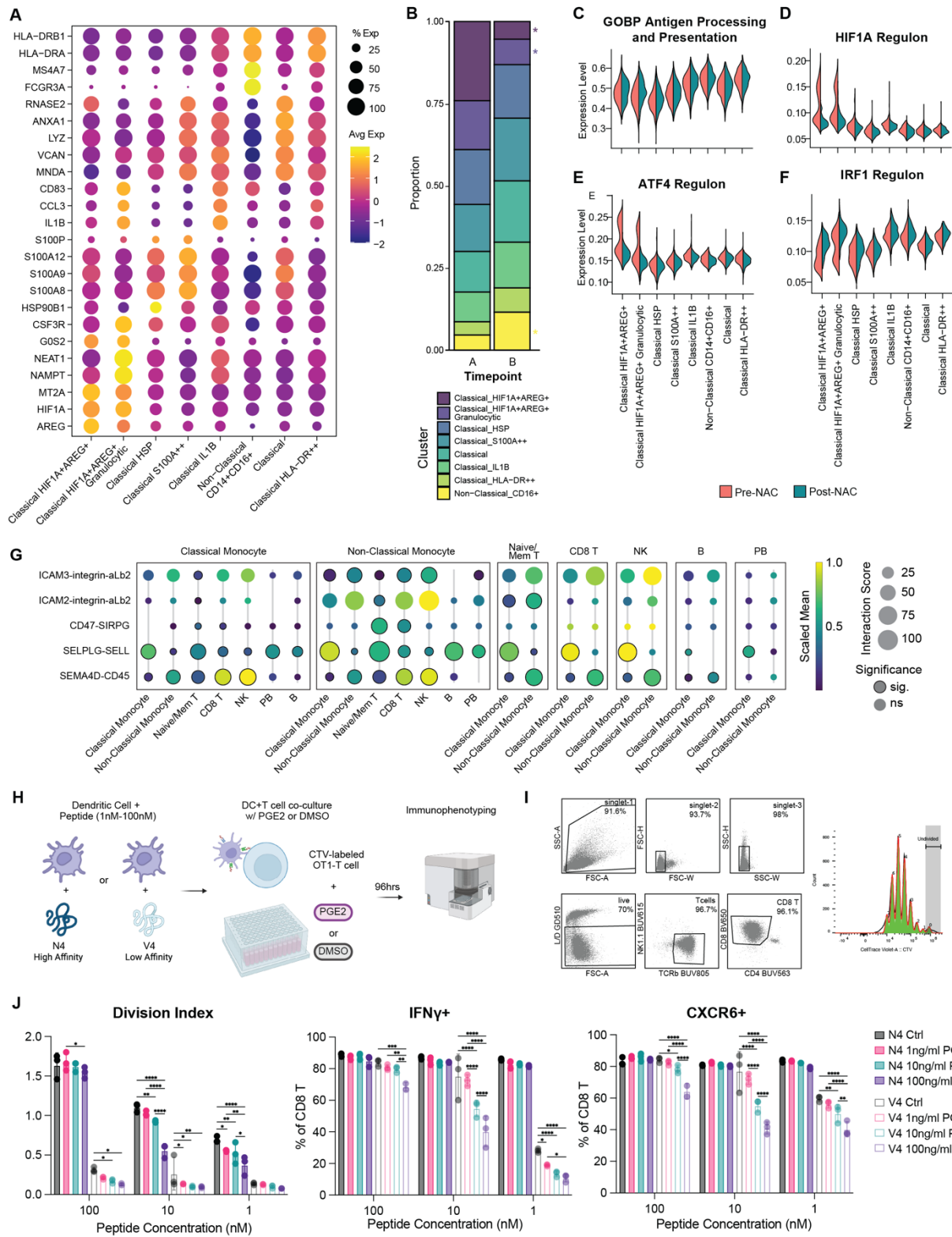

K

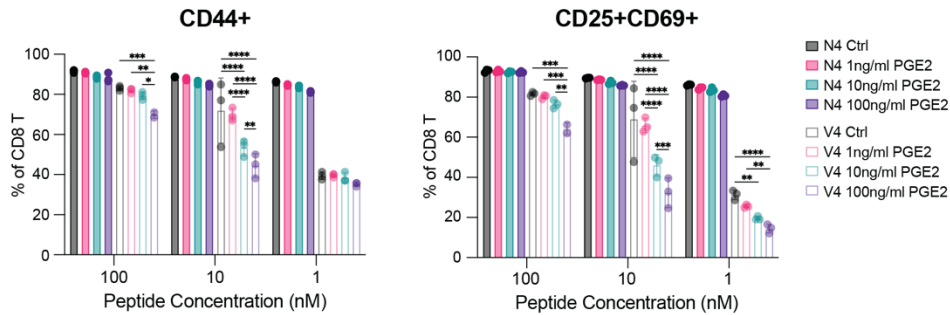

**Supplementary Figure 3. N-Acetylcysteine rescues antigen presentation capacity in disease-severity associated monocytes.** (A) Dot plot of scaled mean expression of marker genes per cluster in the monocyte subset. (B) Stacked bar plot of myeloid cell type proportion changes averaged across samples stratified by Pre-NAC (A) or Post-NAC (B). Differences that are significantly different between timepoints by scCODA are marked with an asterisk. (C-F) Violin plot of single-cell GOBP Antigen Processing and Presentation of Exogenous Antigen pathway normalized enrichment scores (Enrichr) (C), HIF1A Regulon (SCENIC) expression (D), ATF4 Regulon (SCENIC) expression (E), or IRF1 Regulon (SCENIC) expression (F), split by timepoint within each monocyte subcluster. (G) Dot plot depicting scaled mean expression of selected interacting partners per cell-type pair using CellPhoneDBv5 on on-treatment samples. (H) Schematic of experimental design to determine the impact of PGE2 on CD8+ T cell priming. (I) Representative flow cytometry gating to isolate CD8+ T cells. (Left). Representative quantification of CD8+ T cell generations by CTV labeling (Proliferation platform, FlowJo) (Right). (J,K) Dose-dependent impact of PGE2 on CD8+ T cell priming. (J) Division index of CD8+ T cells determined by Cell Trace Violet labeling (Left). Quantification of IFNγ positivity in stimulated CD8+ T cells (Middle). Quantification of CXCR6 positivity in stimulated CD8+ T cells (Right). (K) Quantification of CD44 positivity in stimulated CD8+ T cells (Left). Quantification of CD25 and CD69 positivity in stimulated CD8+ T cells (Right). \*$p < .05$ \*\*$p < .01$ \*\*\*$p < .001$. P values determined by unpaired t-test (J,K).

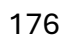

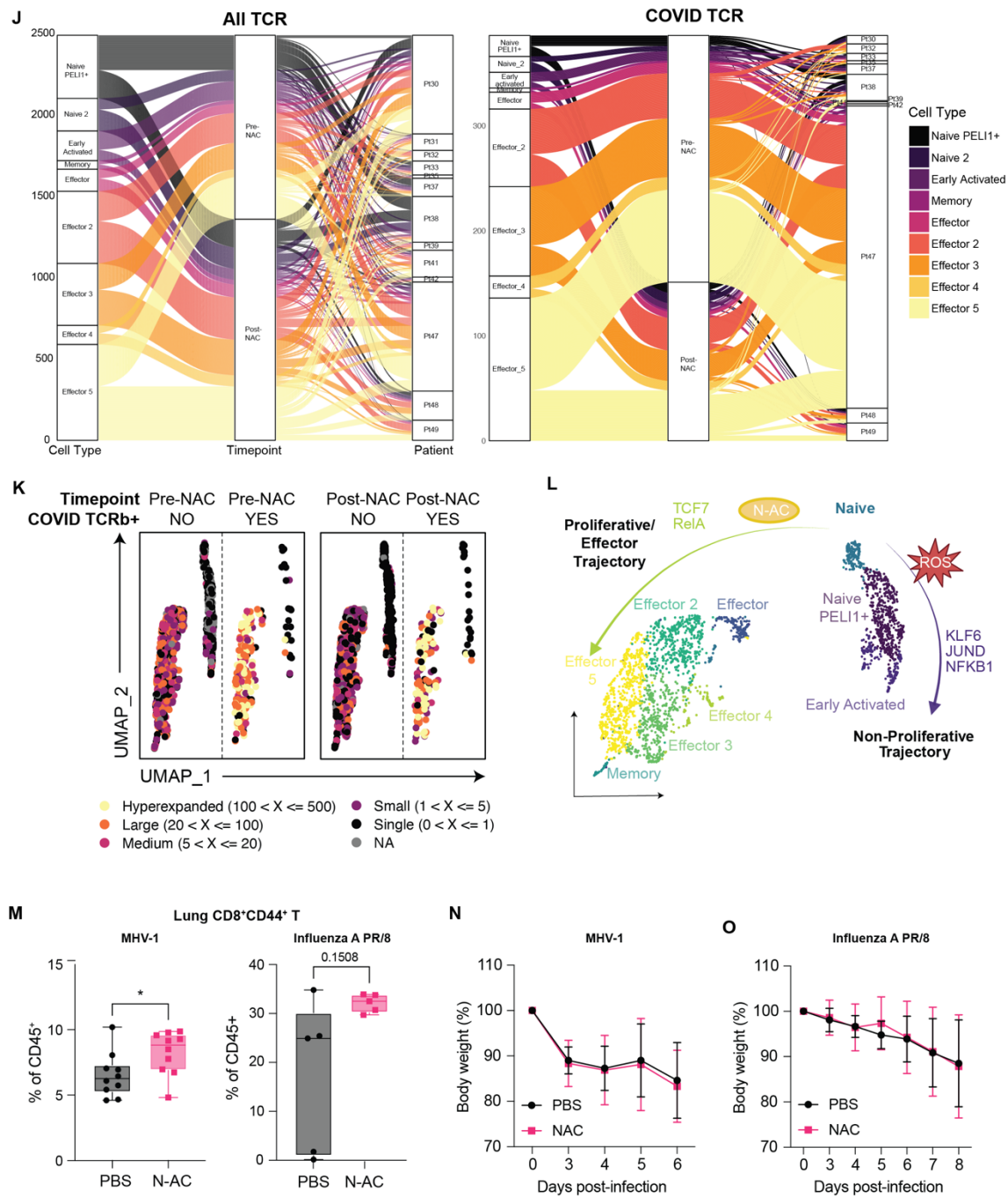

**Supplementary Figure 4. N-Acetylcysteine enhances CD8<sup>+</sup> T activation and effector differentiation *in vitro* and *in vivo*.** (A) Dot plot of scaled mean expression of surface proteins (ADT) per CD8<sup>+</sup> T cell subcluster. (B) Dot plot of scaled mean expression of marker genes per CD8<sup>+</sup> T cell subcluster. (C) Dot plot of scaled mean expression of regulon expression (SCENIC) in CD8<sup>+</sup> T subclusters. (D) Stacked bar plot of CD8<sup>+</sup> T cell type proportion changes averaged across samples stratified by Pre-NAC (A) or Post-NAC (B). Differences that are significantly different between timepoints by scCODA are marked with an asterisk. (E) Feature plot of PELI1 gene expression split by timepoint in CD8<sup>+</sup> T cells. (F) PELI-1 protein expression in primary human T cells activated for 48 hours under normoxic or hypoxic conditions. (G,H) Feature plot

of KLF6 (G) and JUND (H) Regulon expression (SCENIC) split by timepoint in CD8<sup>+</sup> T cells. (I) Clonal diversity quantification by timepoint using clonal diversity function from scRepertoire. (J) Alluvial plot depicting distribution of TCR clones by cell type, timepoint, and patient, either unselected (All TCR, left) or selected for putative COVID-19 reactive clones based on TCRb sequence (COVID TCR, right). (K) Clonal frequency of TCR clones either with or without TCRb sequences that overlap with TCRb sequences identified as COVID-19 reactive in VDJDb. (L) Proposed mechanisms driving canonical and aberrant CD8<sup>+</sup> T cell trajectories in severe COVID-19. (M) Quantified changes in proportions of activated CD44<sup>+</sup>PD1<sup>+</sup>CD8<sup>+</sup> T cells in lung tissue of MHV-1 (left, N=10 per group) or Influenza A PR/8 (right, N=5 per group) infected mice treated with N-AC or PBS control. (N,O) Body weight percentage relative to starting weight at day 0 in mice infected with MHV-1 (N) or Influenza A PR/8 (O), with or without N-AC as indicated. P values calculated using unpaired t-tests (M).

**Supplementary Table 1.** Characteristics of INCOV and MSKCC patients in Figure 1.

**Supplementary Table 2.** cNMF protein weights per module.

**Supplementary Table 3.** Statistical Analysis of cNMF protein module scores across groups.

**Supplementary Table 4.** cNMF metabolite weights per module.

**Supplementary Table 5.** Statistical Analysis of cNMF metabolite module scores across groups.

**Supplementary Table 6.** Clinical characteristic of patients treated on NCT04374461 as well as historical controls.

**Supplementary Table 7.** Plasma metabolites Post-NAC vs Pre-NAC.

**Supplementary Table 8.** Plasma Olink proteomics Post-NAC vs Pre-NAC.

**Supplementary Table 9.** Cell type composition of PBMC object.

**Supplementary Table 10.** Cell type composition of monocyte object.

**Supplementary Table 11.** DEG analysis of monocytes.

**Supplementary Table 12.** CellPhoneDB dominant interactions from on-treatment PBMCs.

**Supplementary Table 13.** CellPhoneDB dominant interactions from pre-treatment PBMCs.

**Supplementary Table 14.** Cell type composition of CD8<sup>+</sup> T cell object.

**Supplementary Table 15.** DEG analysis of CD8<sup>+</sup> T cells.

**Supplementary Table 16.** Differentially expressed regulons with treatment per CD8<sup>+</sup> T cell cluster.
